# GroceryDB: Prevalence of Processed Food in Grocery Stores

**DOI:** 10.1101/2022.04.23.22274217

**Authors:** Babak Ravandi, Gordana Ispirova, Michael Sebek, Peter Mehler, Albert-László Barabási, Giulia Menichetti

**Author notes:** These authors contributed equally (Babak Ravandi, Gordana Ispirova, Michael Sebek, Giulia Menichetti).

## Abstract

The offering of grocery stores is a strong driver of consumer decisions, shaping their diet and long-term health. While highly processed food like packaged products, processed meat, and sweetened soft drinks have been increasingly associated with unhealthy diet, information on the degree of processing characterizing an item in a store is not straight forward to obtain, limiting the ability of individuals to make informed choices. Here we introduce GroceryDB, a database with over 50,000 food items sold by Walmart, Target, and Wholefoods, unveiling how big data can be harnessed to empower consumers and policymakers with systematic access to the degree of processing of the foods they select, and the potential alternatives in the surrounding food environment. The extensive data gathered on ingredient lists and nutrition facts enables a large-scale analysis of ingredient patterns and degrees of processing, categorized by store, food category, and price range. Our findings reveal that the degree of food processing varies significantly across different food categories and grocery stores. Furthermore, this data allows us to quantify the individual contribution of over 1,000 ingredients to ultra-processing. GroceryDB and the associated http://TrueFood.Tech/ website make this information accessible, guiding consumers toward less processed food choices while assisting policymakers in reforming the food supply.

## Introduction

Food ultra-processing has drastically increased productivity and shelf-time, addressing the issue of food availability to the detriment of food systems sustainability and health [1–4]. Indeed, there is increasing evidence that our over-reliance on ultra-processed food (UPF) has fostered unhealthy diet [5]. The sheer number of peer-reviewed articles investigating the link between the degree of food processing and health embodies a general consensus among independent researchers on the health relevance of UPF, contributing up to 60% of consumed calories in developed nations [6–8]. For instance, recent studies have linked the consumption of UPF to non-communicable diseases like metabolic syndrome [9–15], and exposure to industrialized preservatives and pesticides [16–20]. This body of work has driven a paradigm shift from focusing solely on food security, which emphasizes access to affordable food, to prioritizing nutrition security [21, 22]. Nutrition security stresses equitable access to healthy, safe, and affordable foods essential for optimal health and well-being, as defined by the USDA [23, 24], echoing the recent White House Conference on Hunger, Nutrition, and Health [25].

Much of UPF reaches consumers through grocery stores, as documented by the National Health and Nutrition Examination Survey (NHANES), indicating that in the US over 60% of the food consumed comes from grocery stores (Figure S1). The high reliance on UPFs and their potential negative health effects raise numerous critical questions, such as: 1) How can we determine the degree of processing of food items? 2) What methods can be used to quantify the extent of food processing in the food supply? 3) What alternatives can we identify to reduce UPF consumption?

Measuring the degree of food processing is a key step in addressing these questions, but it is not straightforward. Indeed, food labels often display mixed messages, partly driven by reductionist metrics focusing on one nutrient at a time [26], and partly because of the contrasting criteria on how to classify processed foods [27]. The ambiguity and inconsistency of current food processing classification systems (FPCS) have led to conflicting results on their role as risk factors for non-communicable chronic diseases [28, 29]. Some of these classification systems also suffer from poor inter-rater reliability and lack of reproducibility, issues rooted in purely descriptive expertise-based approaches, leaving room for ambiguity and differences in interpretation [27, 28, 30]. Hence, there is a growing call among scientists for a more objective definition of the degree of food processing, based on underlying biological mechanisms rather than subjective opinions of different research groups [28]. Among the proposed areas for aligning food processing definitions, the nutritional profile of food is currently the only aspect consistently regulated and reported worldwide [27, 28, 31].

The research efforts outlined in [28] align with a growing demand for high-quality and internationally comparable statistics to promote objective metrics, reproducibility, and data-driven decision-making, advancing our convergence towards the Sustainable Development Goals (SDGs) [32, 33]. Artificial intelligence (AI) methodologies [33–36], in particular, are increasingly being utilized for their potential as objective, data-driven tools to advance populations’ nutrition security, a concept underpinning SDGs ‘zero-hunger’, ‘good health and well-being’, ‘industry, innovation, and infrastructure’, and ‘reduce inequalities’.

Responding to the need for objective and scalable metrics to ensure nutrition security, we have recently harnessed machine learning (ML) to create and fully automate our Food Processing Score (FPro) [37]. FPro is a continuous index derived by training an ML model to predict manual labels of processing techniques based on the overall nutrient profile of a food (see Methods and Section S4). To teach our algorithm how to score processing from nutrients, we leveraged the labels provided by NOVA, currently the most widely used system to classify foods according to processing-related criteria, offering us an extensive array of epidemiological literature for comparative analysis [38, 39]. However, the FPro algorithm can accommodate different FPCS such as EPIC [40], UNC [41], or SIGA [42]. We rigorously tested the predictive power of FPro for epidemiological outcomes with an Environment-Wide Association Study (EWAS), leveraging multiple cycles of USDA’s model food databases and national food consumption surveys [37].

Here, building on the versatility and scalability of the FPro algorithm, we extend our analysis beyond “model foods” tailored for epidemiological databases and instead analyze real-world data encompassing over 50,000 products obtained from major US grocery store websites. This extensive dataset underpins the development of GroceryDB, an open-source database of foods and beverages, featuring comprehensive metadata on nutritional content, ingredient list, and price for each item, collected from publicly available online markets of Walmart, Target, and Whole Foods. Our objective is to demonstrate how ML can effectively analyze large-scale real-world food composition data, and translate this wealth of information into the degree of processing for any food in grocery stores, facilitating consumer decision-making and informing public health initiatives aimed at enhancing the overall quality of the food environment. GroceryDB, accessible to the public at http://TrueFood.Tech/, offers both the data and methodologies needed to quantify food processing and analyze the structure of ingredients within the U.S. food supply. This initiative not only lays the groundwork for similar efforts globally, aimed at promoting better-informed dietary choices, but also underscores the critical role of open-access, internationally comparable data in advancing global nutrition security.

## Main

For each food, we automated the process of determining the extent of food processing using FPro, which translates the nutritional content of a food item, as reported by the nutrition facts, into its degree of processing [37]. In Figure 1, we illustrate the use of FPro by offering the processing score of three products in the breads and yogurt categories, allowing us to compare their degree of processing. Indeed, the Manna Organics multigrain bread is made from whole wheat kernels, barley, and rice without additives, added salt, oil, and even yeast, resulting in a low processing score of *FPro* = 0.314. However, the Aunt Millie’s and Pepperidge Farmhouse breads include ‘resistant corn starch’, ‘soluble corn fiber’, and ‘oat fiber’, requiring additional processing to extract starch and fiber from corn and oat to be used as an independent ingredient (Figure 1a), resulting in much higher processing score of *FPro* = 0.732 and *FPro* = 0.997. Similarly, the Seven Stars Farm yogurt (*FPro* = 0.355) is a whole milk yogurt made from ‘grade A pasteurized organic milk’, yet the Siggi’s yogurt (*FPro* = 0.436) uses ‘Pasteurized Skim Milk’ that requires more processing to obtain 0% fat. Finally, the Chobani Cookies & Cream yogurt relies on cane sugar as the second most dominant ingredient, and on a cocktails of additives like ‘caramel color’, ‘fruit pectin’, and ‘vanilla bean powder’ making it a highly processed yogurt, resulting in a high processing score *FPro* = 0.918.

**Figure 1:**
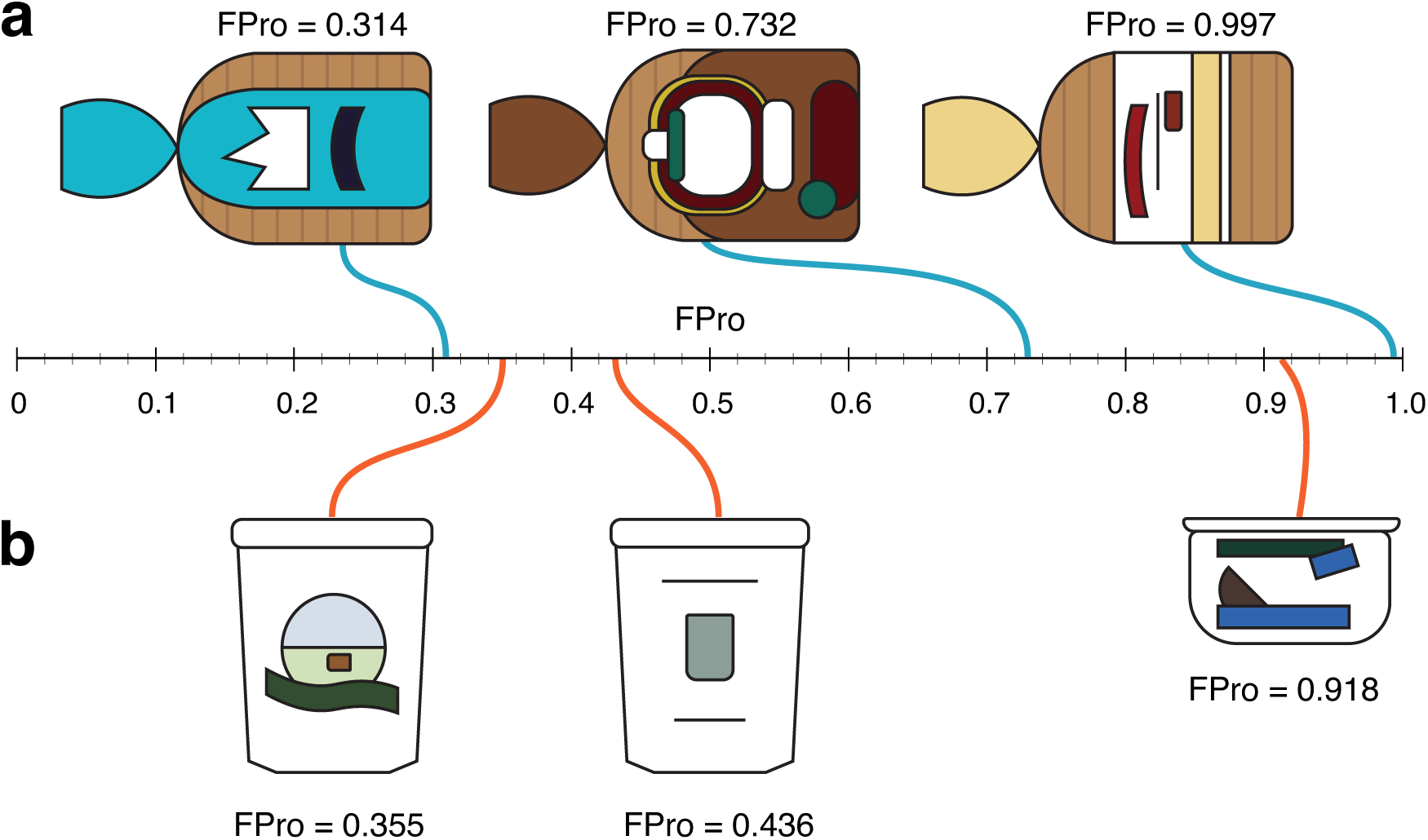
Degrees of Food Processing in Three Categories. FPro allows us to assess the extent of food processing in three major US grocery stores, and it is best suited to rank foods within the same category. **(a)** In breads, the Manna Organics multi-grain bread, offered by WholeFoods, is mainly made from ‘whole wheat kernels’, barley, and brown rice without any additives, added salt, oil, and yeast, with *FPro* = 0.314. However, the Aunt Millie’s (*FPro* = 0.732) and Pepperidge Farmhouse (*FPro* = 0.997) breads, found in Target and Walmart, include ‘soluble corn fiber’ and ‘oat fiber’ with additives like ‘sugar’, ‘resistant corn starch’, ‘wheat gluten’, and ‘monocalcium phosphate’. **(b)** The Seven Stars Farm yogurt (*FPro* = 0.355) is made from the ‘grade A pasteurized organic milk’. The Siggi’s yogurt (*FPro* = 0.436) declares ‘Pasteurized Skim Milk’ as the main ingredients that has 0% fat milk, requiring more food processing to eliminate fat. Lastly, the Chobani Cookies & Cream yogurt (*FPro* = 0.918) has cane sugar as the second most dominant ingredient combined with multiple additives like ‘caramel color’, ‘fruit pectin’, and ‘vanilla bean powder’, making it a highly processed yogurt.

We assigned an FPro score to each food in GroceryDB by leveraging our ML classifier FoodProX, which takes as input the mandatory information captured by the nutrition facts (Methods). We find that the distribution of the FPro scores in the three stores is rather similar: in each store, we observe a monotonically increasing curve (Figure 2a), indicating that minimally-processed products (low FPro) represent a relatively small fraction of the inventory of grocery stores, the majority of the offerings being in the ultraprocessed category (high FPro). Although less-processed items make up a smaller share of the overall inventory, they likely account for a proportionally larger portion of actual purchases, highlighting a discrepancy between sales data and available food options. Nevertheless, we identified systematic differences between stores: Whole Foods offers a greater selection of minimally processed items and fewer ultra-processed options, whereas Target has a particularly high proportion of ultra-processed products (high FPro).

**Figure 2:**
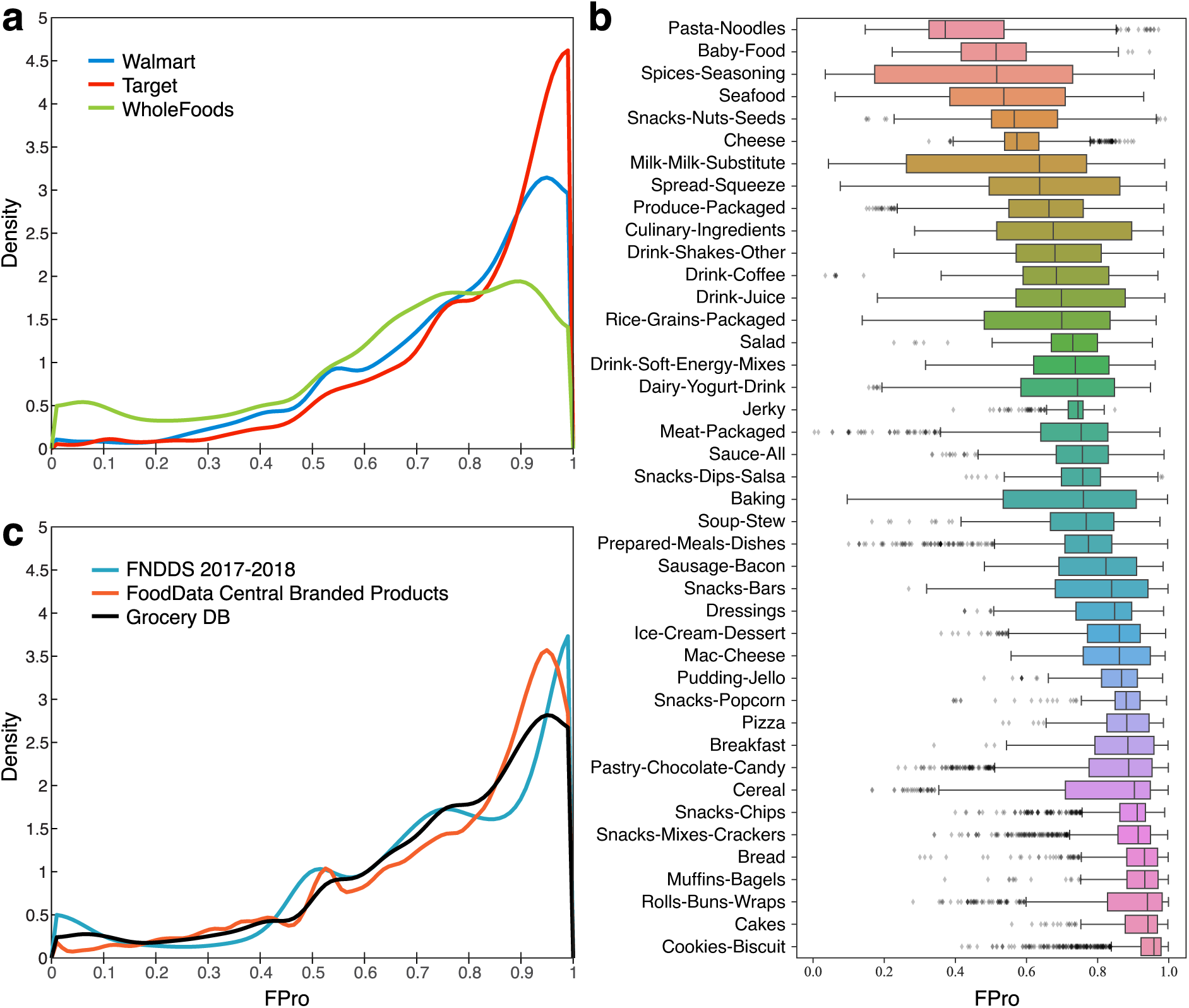
Food Processing in Grocery Stores. (**a**) The distribution of FPro scores from the three stores follows a similar trend, a monotonically increasing curve, indicating that the number of low FPro items (unprocessed and minimally-processed) offered by the grocery stores is relatively lower than the number of high FPro items (highly-processed and ultra-processed items), and the majority of offerings are ultra-processed (Methods for FPro calculation). (**b**) Distribution of FPro scores for different categories of GroceryDB. The distributions indicate that FPro has a remarkable variability within each food category, confirming the different degrees of food processing offered by the stores. Unprocessed foods like eggs, fresh produce, and raw meat are excluded (Section S7). (**c**) The distributions of FPro scores in GroceryDB compared to two USDA nationally representative food databases: the USDA Food and Nutrient Database for Dietary Studies (FNDDS) and FoodData Central Branded Products (BFPD). The similarity between the distributions of FPro scores in GroceryDB, BFPD, and FNDDS suggests that GroceryDB offers a comprehensive coverage of foods and beverages (Section S6).

FPro also captures the inherent variability in the degree of processing per food category. As illustrated in Figure 2b, we find a small variability of FPro scores in categories like jerky, popcorn, chips, bread, biscuits, and mac & cheese, indicating that consumers have limited choices in terms of degree of processing in these categories (Section S7 for harmonizing categories between stores). Yet, in categories like cereals, milk & milk-substitute, pasta-noodles, and snack bars, FPro varies widely, reflecting a wider extent of possible choices from a food processing perspective.

We compared the distribution of FPro in GroceryDB with the latest USDA Food and Nutrient Database for Dietary Studies (FNDDS), offering a representative sample of the consumed food supply (Figure 2c). The similarity between the distributions of FPro scores obtained from GroceryDB and FNDDS suggests that GroceryDB also offers a representative sample of foods and beverages in the supply chain. Additionally, we compared GroceryDB with the USDA Global Branded Food Products Database (BFPD), which contains 1,142,610 branded products, finding that the distributions of FPro in GroceryDB and BFPD follow similar trends (Figure 2c). While BFPD contains 22 times more foods than GroceryDB, only an estimated 44% of GroceryDB’s products are represented in BFPD, even after accounting for potential variability in food names and ingredient lists (Section S6). This indicates that while BFPD offers an extensive representation of branded products, it does not fully capture the current offering of stores. Furthermore, we compared GroceryDB with Open Food Facts (OFF) [43], another extensive collection of branded products collected through crowd-sourcing, containing 426,000 products with English ingredient lists. We find that less than 40% of the products in GroceryDB are present in OFF (Figure S4), a small overlap, suggesting that monitoring the products currently offered in grocery stores may provide a more accurate account of the food supply available to consumers.

### Food Processing and Caloric Intake

The depth and the resolution of the data collected in GroceryDB allow us to unveil some of the complexity regarding the relation between price and calories. Among all categories in GroceryDB, a 10% increase in FPro results in 8.7% decrease in the price per calorie of products, as captured by the dashed line in Figure 3A. However, the relationship between FPro and price per calorie strongly depends on the food category (Section S8). For example, in soups & stews the price per calorie drops by 24.3% for 10% increase in FPro (Figure 3b), a trend observed also in cakes, mac & cheese, and ice cream (Figure S8). This means that on average, the most processed soups & stews, with *FPro* ≈ 1, are 67.72% cheaper per calories than the minimally-processed alternatives with *FPro* ≈ 0.4 (Figure 3e). In contrast, in cereals price per calorie drops only by 1.2% for 10% increase in FPro (Figure 3c), a slow decrease observed also for seafood and yogurt products (Figure S8). Interestingly, we find an increasing trend between FPro and price in the milk & milk-substitute category (Figure 3d), partially explained by the higher price of plant-based milk substitutes, that require more extensive processing than the dairy-based milks.

**Figure 3:**
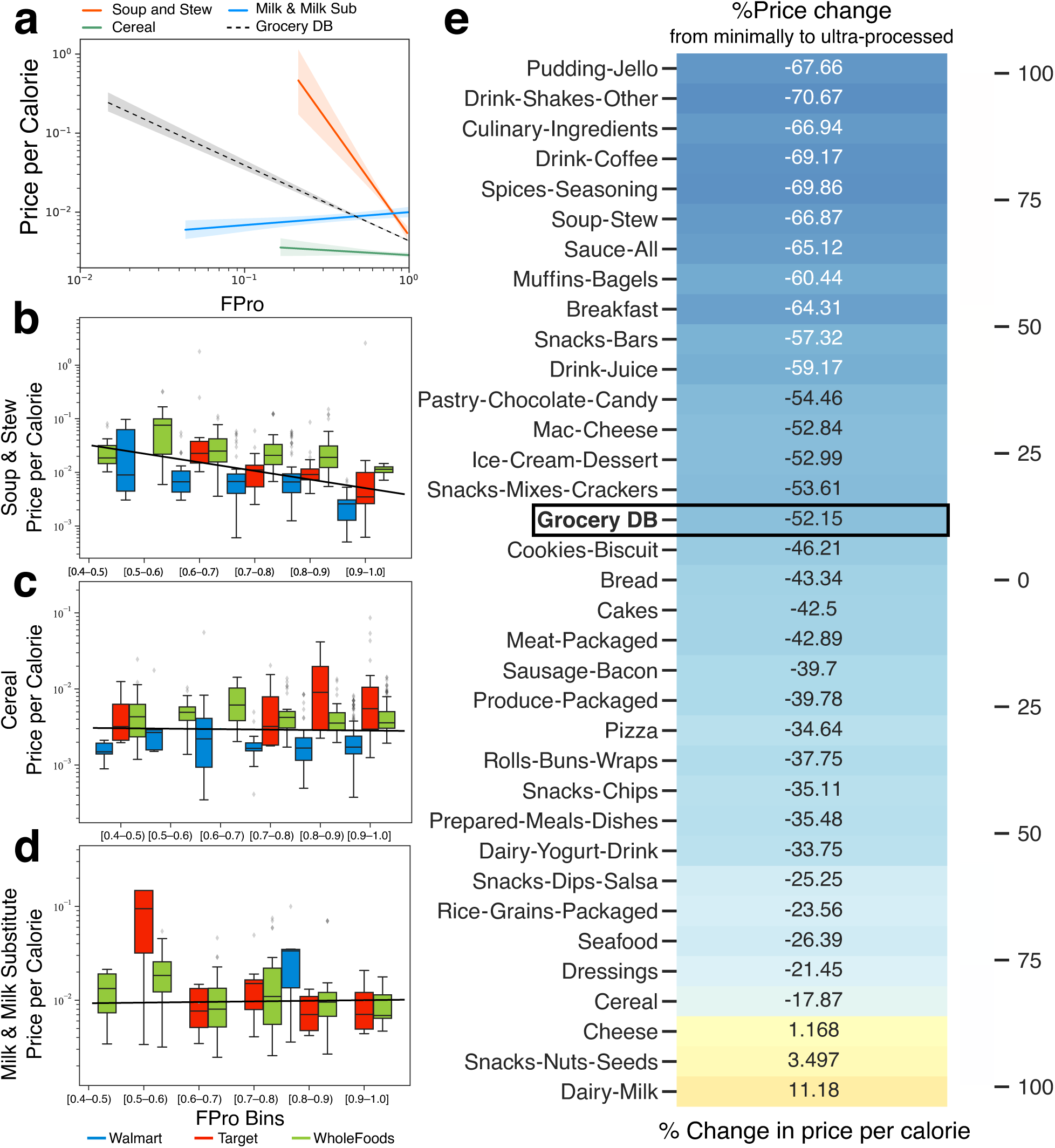
Price and Food Processing. **(a)** Using robust linear models, we assessed the relationship between price and food processing (Figure S8 for regression coefficients of all categories). We find that price per calories drops by 24.3% and 1.2% for 10% increase in FPro in soup & stew and cereals, respectively. Also, we observe a 8.7% decrease across all foods in GroceryDB for 10% increase in FPro. Interestingly, in milk & milk-substitute, price per calorie increases by 1.6% for 10% increase in FPro, partially explained by the higher price of plant-based milks that are more processed than regular dairy milk. **(bd)** Distributions of price per calorie in the linear bins of FPro scores for each store (Figure S7 illustrates the correlation between price and FPro for all categories). In soup & stew, we find a steep decreasing slope between FPro and price per calorie, while in cereals we observe a smaller effect. In milk & and milk-substitute, price tends to slightly increase with higher values of FPro. **(e)** Percentage of change in price per calorie from the minimally-processed products to ultra-processed products in different food categories. This analysis was performed by comparing the average price per calorie of the top 10% most processed items with the top 10% least processed items within each category. In the full GroceryDB, on average, the ultra-processed items are 52.09% cheaper than their minimally-processed alternatives.

### Choice Availability and Food Processing

Not surprisingly, GroceryDB documents differences in the offering of the three stores we analyzed: while WholeFoods offers a selection of cereals with a wide range of processing levels, from minimally-processed to ultra-processed, in Walmart the available cereals are limited to products with higher FPro values (Figure 4a). To understand the roots of these differences, we investigated the ingredients of cereals offered by each grocery store, one of the most popular staple crops, consumed by 283 million Americans in 2020 [44]. We find that cereals offered by WholeFoods rely on less sugar, less natural flavors, and added vitamins (Figure 4b). In contrast, cereals in Target and Walmart tend to contain corn syrup, a sweetener associated with enhanced absorption of dietary fat and weight gain [45]. Corn syrup is largely absent in the WholeFoods cereals, partially explaining the wider range of processing scores characterising cereals offered by the store (Figure 4a).

**Figure 4:**
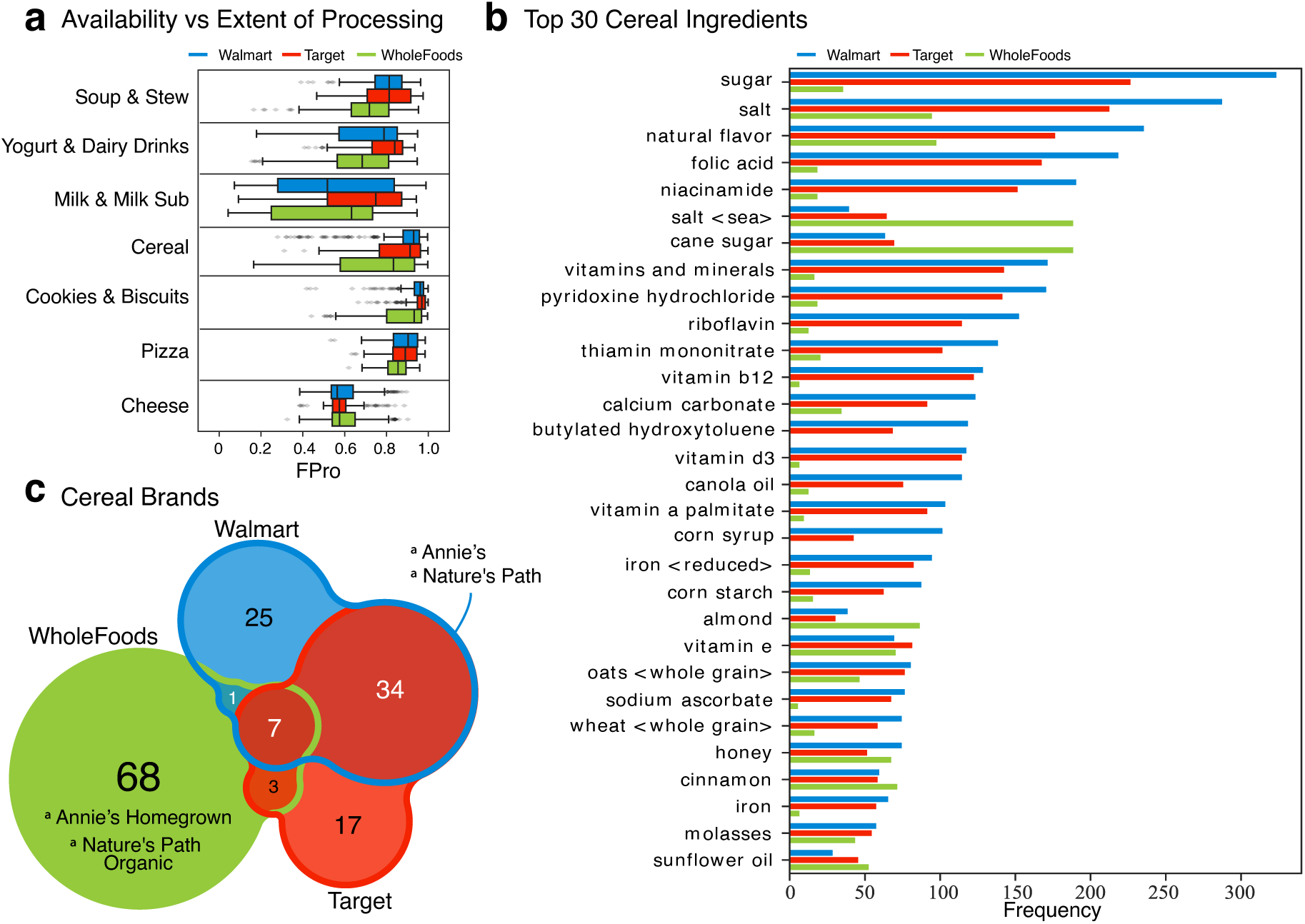
The Difference between Stores in Term of Processing. The degree of processing of food choices depends on the grocery store and food category. (**a**) The degree of processing of food items offered in grocery stores, stratified by food category. For example, in cereals, WholeFoods shows a higher variability of FPro, implying that consumers have a choice between low and high processed cereals. Yet, in pizzas all supermarkets offer choices characterised by high FPro values. Lastly, all cheese products are minimally-processed, showing consistency across different grocery stores. (**b**) The top 30 most reported ingredients in cereals shows that WholeFoods tends to eliminate corn syrup, uses more sunflower oil and less canola oil, and relies less on vitamin fortification. In total, GroceryDB has 1,168 cereals from which 973 have ingredient lists. 309, 260, and 395 cereals are from Walmart, Target, and WholeFoods, respectively. (**c**) The brands of cereals offered in stores partially explains the different patterns of ingredients and variation of FPro. While Walmart and Target have a larger intersection in the brands of their cereals, WholeFoods tends to supply cereals from brands not available elsewhere.

The brands offered by each store could also help explain the different patterns. We found that while Walmart and Target have a large overlap in the list of brands they carry, WholeFoods relies on different suppliers (Figure 4c), largely unavailable in other grocery stores. In general, WholeFoods offers less processed soups & stews, yogurt & yogurt drinks, and milk & milk-substitute (Figure 4a). In these categories Walmart’s and Target’s offerings are limited to higher FPro values. Lastly, some food categories like pizza, mac & cheese, and popcorn are highly processed in all stores (Figure 4a). Indeed, pizzas offered in all three chains are limited to high FPro values, partially explained by the reliance on substitute ingredients like “imitation mozzarella cheese,” instead of “mozzarella cheese”.

While grocery stores offer a large variety of products, the offered processing choices can be identical in multiple stores. For example, GroceryDB has a comparable number of cookies & biscuits in each chain, with 453, 373, and 402 items in Walmart, Target, and WholeFoods, respectively. The degree of processing of cookies & biscuits in Walmart and Target are nearly identical (0.88 *< FPro <* 1), limiting consumer nutritional choices in a narrow range of processing (Figure 4a). In contrast, WholeFoods not only offers a large number of items (402 cookies & biscuits), but it also offers a wider choices of processing (0.57 *< FPro <* 1)

### Organization of Ingredients in the Food Supply

Food and beverage companies are required to report the list of ingredients in the descending order of the amount used in the final product. When an ingredient itself is a composite, consisting of two or more ingredients, FDA mandates parentheses to declare the corresponding sub-ingredients (Figure 5a-b) [46]. We organized the ingredient list as a tree (Methods), allowing us to compare a highly processed cheesecake with a less processed alternative (Figure 5). In general, we find that products with complex ingredient trees are more processed than products with simpler and fewer ingredients (Section S9.3). For example, the ultra-processed cheesecake in Figure 5a has 43 ingredients, 26 additives, and 3 branches with sub-ingredients. In contrast, the minimally-processed cheesecake has only 14 ingredients, 5 additives, and 2 branch with sub-ingredients (Figure 5b). As illustrated by the cheesecakes example, ingredients used in the food supply are not equally processed, prompting us to ask: which ingredients contribute the most to the degree of processing of a product? To answer this we introduce the Ingredient Processing Score (IgFPro), defined as

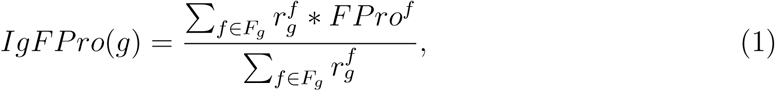

**Figure 5:**
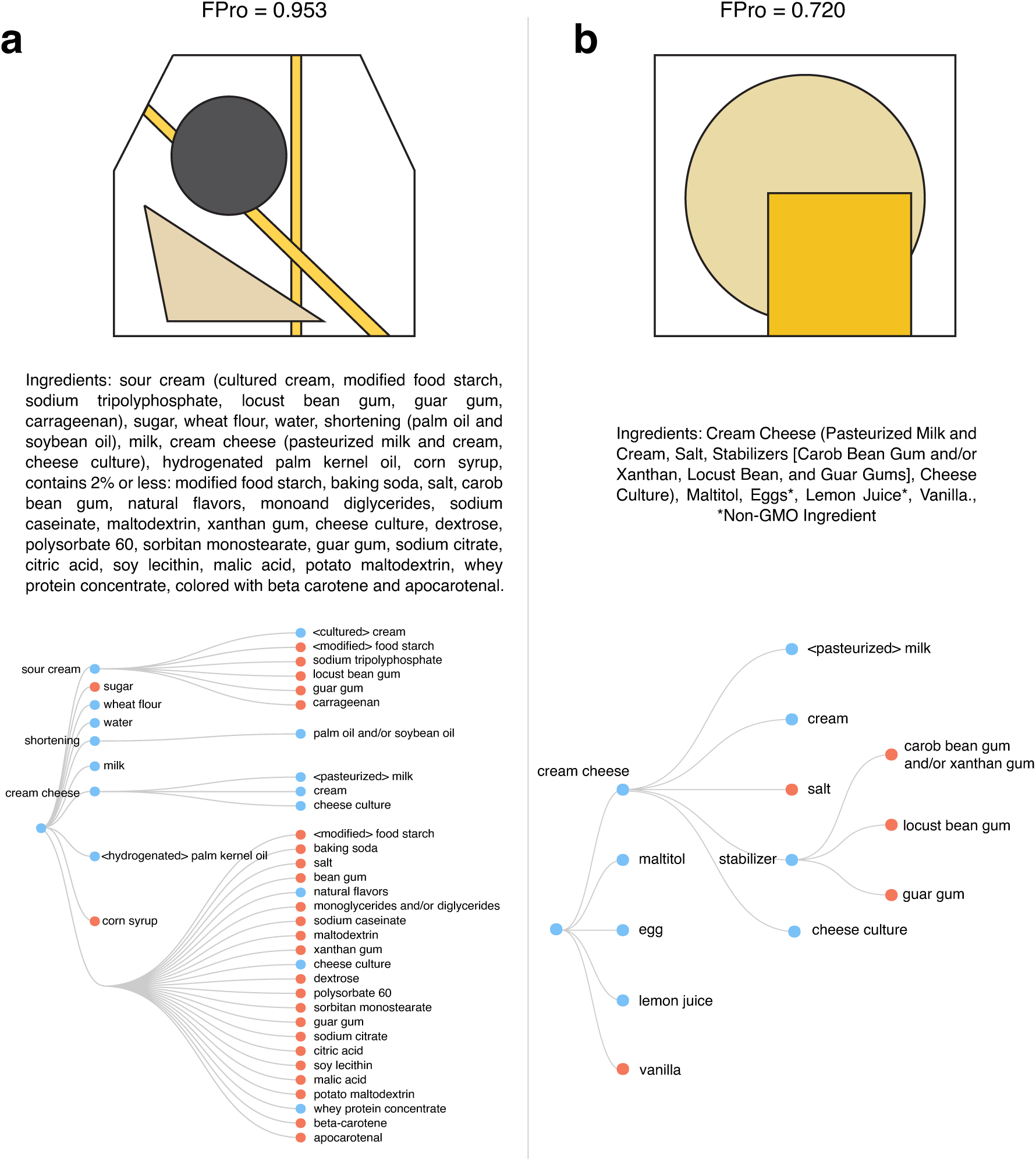
Ingredient Trees. GroceryDB organizes the ingredient list of products into structured trees, where the additives are marked as orange nodes (Methods and Section S9). **(a)** Edwards Desserts Original Whipped Cheesecake is a highly processed cheesecake that contains 43 ingredients from which 26 are additives, resulting in a complex ingredient tree with 3 branches of sub-ingredients. **(b)** Pearl River Mini No Sugar Added Chessecake is a minimally-processed cheesecake that has a simpler ingredient tree with 14 ingredients, 5 additives, and 2 branches with sub-ingredients.

where 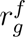 ranks an ingredient *g* in decreasing order based on its position in the ingredient list of each food *f* that contains *g* (Section S9.5). IgFPro ranges between 0 (unprocessed) and 1 (ultra-processed), allowing us to rank-order ingredients based on their contribution to the degree of processing of the final product. We find that not all additives contribute equally to ultra-processing. For example, the ultra-processed cheesecake (Figure 5a) has polysorbate 60 (an emulsifier used in cakes for increased volume and fine grain with *IgFPro* = 0.908), and corn syrup (a corn sweetener with *IgFPro* = 0.905) [47], each of which emerging as signals of ultra-processing with high IgFPro scores. In contrast, both the minimally-processed and ultra-processed cheesecakes (Figure 5) contain xanthan gum (*IgFPro* = 0.818), guar gum (*IgFPro* = 0.801), locust bean gum (*IgFPro* = 0.786), and salt (*IgFPro* = 0.777). Indeed, the European Food Safety Authority (EFSA) reported that xanthan gum as a food additive does not pose any safety concern for the general population, and FDA classified guar gum and locust bean gum as generally recognized safe [47].

By the same token, we looked into the oils used as ingredients in branded products to assess which oils contribute the most to UPFs. IgFPro identifies brain octane oil (*IgFPro* = 0.573), flax seed oil (*IgFPro* = 0.69), and olive oil (*IgFPro* = 0.722) as the highest quality oils, having the smallest contribution to ultra-processing. In contrast, palm oil (*IgFPro* = 0.888), vegetable oil (*IgFPro* = 0.866), and soy bean oil (*IgFPro* = 0.862) represent strong signals of ultra-processing (Figure 6a). Indeed, flax seed oil is high in omega-3 fatty acids with several health benefits [48]. In contrast, the blending of vegetable oils, a signature of UPF, is one of the simplest methods to create products with desired texture, stability, and nutritional properties [49].

**Figure 6:**
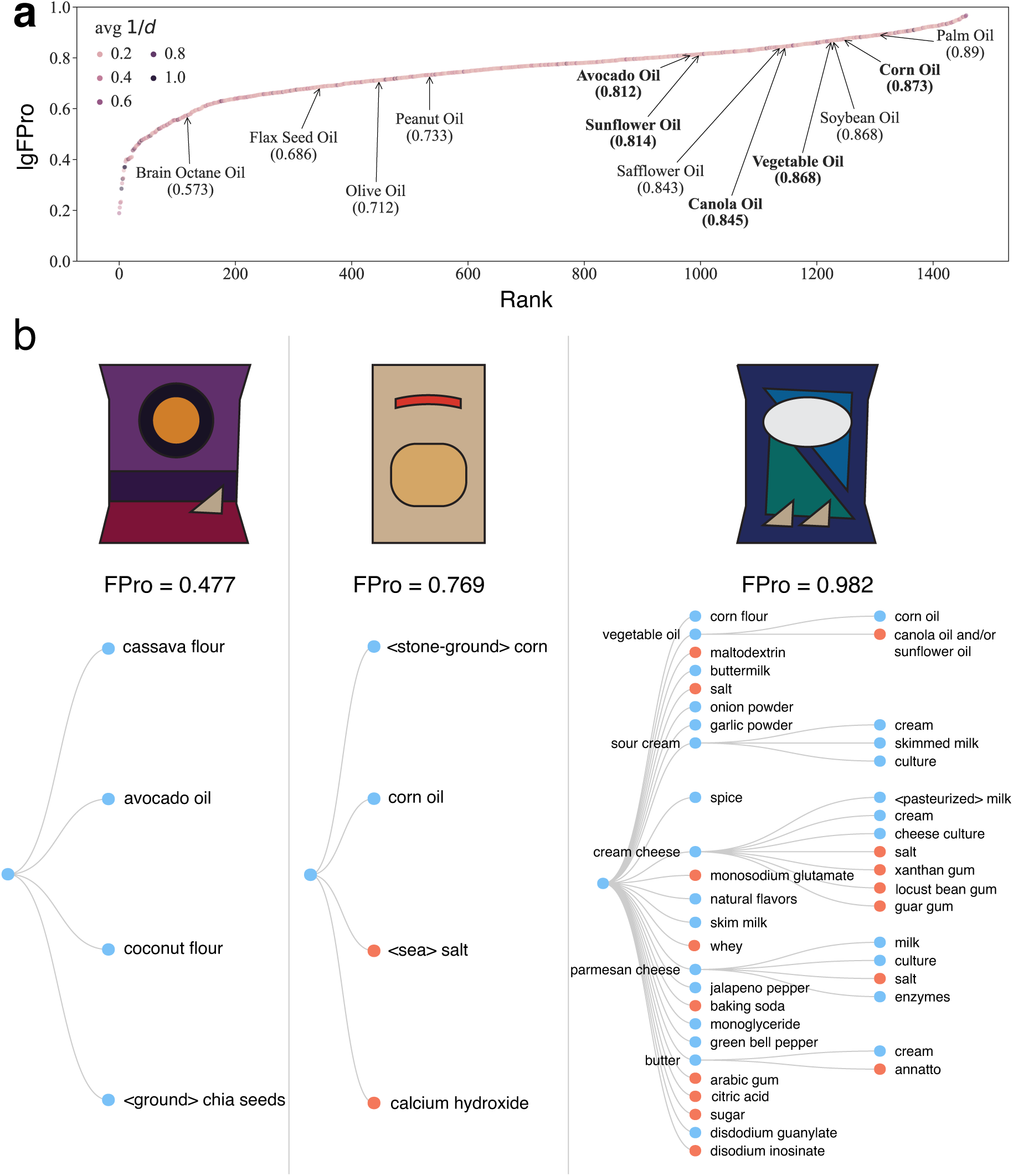
Ingredient Processing Score (IgFPro). To investigate which ingredients contribute most to ultra-processed products, we extend FPro to ingredient lists using Eq. 1. With the introduction of IgFPro, we can rank over 12,000 ingredients by their prevalence and contribution to ultra-processed products prioritizing ingredients and food groups for targeted intervention. A total of 1,676 ingredients are in more than 10 products. **(a)** The IgFPro of all ingredients that appeared in at least 10 products are calculated, rank-ordering ingredients based on their contribution to UPFs. The ingredients are colored based on their distance to the root node, *d*, of the ingredient tree (Methods). The popular oils used as an ingredient are highlighted, with the brain octane, flax seed, and olive oils contributing the least to ultra-processed products. In contrast, the palm, vegetable, and soybean oils contribute the most to ultra-processed products (Section S9.5). **(b)** The patterns of ingredients in the least-processed tortilla chips vs. the ultra-processed tortilla chips. The bold fonts track the IgFPro of the oils used in the three tortilla chips. The minimally-processed Siete tortilla chips (*FPro* = 0.477) uses avocado oil (*IgFPro* = 0.822), and the more processed El Milagro tortilla (*FPro* = 0.769) has corn oil (*IgFPro* = 0.886). In contrast, the ultra-processed Doritos (*FPro* = 0.982) relies on a blend of vegetable oils (*IgFPro* = 0.866), and is accompanied with a much more complex ingredient tree, indicating that there is no single ingredient “bio-marker” for UPFs.

Finally, to illustrate the ingredient patterns characterising UPFs in Figure 6b, we show three tortilla chips, ranked from the “minimally-processed” to the ultra-processed. Relative to the snack-chips category, Siete tortilla is minimally-processed (*FPro* = 0.477), made with avocado oil and blend of cassava and coconut flours. The more processed El Milagro tortilla (*FPro* = 0.769) is cooked with corn oil, grounded corn, and has calcium hydroxide, generally recognized as a safe additive made by adding water to calcium oxide (lime) to promote dispersion of ingredients [47]. In contrast, the ultra-processed Doritos (*FPro* = 0.982) have corn flour, a blend of vegetable oils, and rely on 12 additives to ensure a palatable taste and the texture of the tortilla chip, demonstrating the complex patterns of ingredients and additives needed for ultra-processing (Figure 6b).

In summary, complex ingredient patterns accompany the production of UPFs (Section S9.4). IgFPro captures the role of individual ingredients in the food supply, enabling us to diagnose the processing characteristics of the whole food supply as well as the contribution of individual ingredients.

## Discussion

By combining large-scale data on food composition and ML, GroceryDB uncovers insights on the current state of food processing in the US grocery landscape, enabling us to obtain distributions of food processing scores that capture a remarkable variability in the offerings of different grocery stores. The differences in FPro’s distributions (Figure 2A) indicate that multiple factors drive the range of choices available in grocery stores, from the cost of food and the socio-economic status of the consumers to the distinct declared missions of the supermarket chains: “quality is a state of mind” for WholeFoods Market and “helping people save money so they can live better” for Walmart [50, 51]. Furthermore, the continuous nature of FPro enabled us to conduct a data-driven investigation on the relationship between price and food processing stratified by food category. We find that overall in GroceryDB food processing tends to be associated with the production of more affordable calories, a positive correlation that raises the likelihood of habitual consumption among lower-income populations, ultimately contributing to growing socioeconomic disparities in terms of nutrition security [52–57]. However, it is important to note that the strength and direction of this correlation varies depending on the specific food category under consideration, as exemplified by the opposite trend of milk & milk-substitutes compared to soups & stews (Section S8). Further in-depth analyses are needed to evaluate the effectiveness of intervention strategies targeting specific food groups within diverse food environments.

Governments increasingly acknowledge the impact of processed foods on population health, and its long-term effect on healthcare [58, 59]. For example, the UK spends £18 billion annually on direct medical costs related to non-communicable diseases like obesity [60], while the US incurs $1.1 trillion in yearly food-related human health costs [61,62]. GroceryDB serves as a valuable resource for both consumers and policymakers, offering essential insights to gauge the level of food processing within the food supply. For instance, in categories like cereals, milk & milk alternatives, pasta-noodles, and snack bars, FPro exhibits a wide range, highlighting the substantial variations in the processing levels of products. If consumers had access to this processing data, they could make informed choices, selecting items with significantly different degrees of processing (Figure 2B). Yet, the comprehension of nutrient and ingredient data disclosed on food packaging often poses a challenge to consumers due to unrealistic serving sizes and confusing health claims based on one or a few nutrients. Our primary objective lies in translating this wealth of data into an actionable scoring system, enabling consumers to make healthier food choices and embrace effective dietary substitutions, without overwhelming them with excessive information. Additionally, our approach holds great potential for public health initiatives aimed at improving the overall quality of our food environment, such as strategies reorganizing supermarket layouts, optimizing shelf placements, and thoughtfully designing counter displays [54, 63, 64]. Transforming health-related behaviors is a challenging task [65,66], hence easily adoptable dietary modifications along with environmental nudges could make it easier for individuals to embrace healthier choices.

Currently, FPro partially draws from expertise-based food processing classifications due to limited data concerning compound concentrations indicative of food matrix alterations, such as cellular wall transformations or industrial processing techniques. However, a comprehensive mapping of the “Dark Matter of Nutrition”, encompassing chemical concentrations for additives and processing byproducts, aims to evolve FPro into an unsupervised system, independent of manual classifications [67, 68]. Unlike expertise-based systems, FPro functions as a quantitative algorithm, utilizing standardized inputs to generate reproducible continuous scores, facilitating sensitivity analysis and uncertainty estimations [37] (Section S5). These important features enhance analyses’ reliability, transparency, and interpretability while reducing errors linked to the descriptive nature of manual classifications [28], which have displayed a low degree of consistency among nutrition specialists [69].

The chemical composition of branded products is partially captured by the nutrition facts table and partially reported in the ingredient list, which includes additives like artificial colors, flavors, and emulsifiers. However, comprehensive and internationally well-regulated data on food ingredients is currently limited, as documented by the GS1 UK data crunch analysis which reported an average of 80% inconsistency in products’ data [31], leading us to focus on the nutrition facts to enhance our algorithm’s portability and reproducibility. The nutrition facts alone exhibit excellent performance in discriminating between NOVA classes, confirming how food processing consistently alters nutrient concentrations with reproducible patterns, effectively harnessed by ML [37]. While FPro assesses the degree of food processing by holistically evaluating nutrient concentrations, the few nutrients available on food packaging increase the risk of identifying products with similar nutrition facts but distinct food matrices (e.g., pre-frying, puffing, extrusion-cooking). Indeed, if the chemical panel used to train the algorithm fails to exhaustively capture matrix modifications induced by processing and cooking, FPro and the substitution algorithm implemented at http://TrueFood.Tech/, remain blind to these chemical-physical changes. Incorporating disambiguated ingredients in FPro, such as the ultra-processing markers characterized by SIGA [70], may offer a solution until larger composition tables for branded products become available (Section S5).

In summary, our work represents a departure from traditional food classification systems, advancing toward the use of ML methodologies to model the chemical complexity of food (Section S1). Despite the limited information provided by FDA-regulated nutrition labels, GroceryDB and FPro offer a data-driven approach that enables a substitution algorithm capable of recommending similar but less processed alternatives for any food in GroceryDB. Together, GroceryDB and the TrueFood platform highlight the importance of data transparency in grocery store inventories, a key factor that directly shapes consumer choices.

## Methods

### Data Collection

We compiled publicly accessible data on food products available at Walmart, Target, and Whole Foods through their respective online platforms. Each store organizes its food items hierarchically. Utilizing these categorizations, we systematically navigated through the stores’ websites to identify specific food items. To ensure consistency, we standardized the food category hierarchy within GroceryDB by comparing and aligning the classification systems employed by each store. These stores sourced nutrition facts from physical food labels and provided digital versions for each food item. This data enabled us to standardize nutrient concentrations to a uniform measure of 100 grams and employ FoodProX to evaluate the degree of food processing for each item. Lastly, all data was collected in May 2021.

### Calculation of the Food Processing Score (FPro)

Processing alters the nutrient profile of food, changes that are detectable and categorizable using ML [37, 71, 72]. Hence, we developed FoodProX [37], a random forest classifier that can translate the combinatorial changes in the nutrient amounts induced by food processing into a food processing score (FPro). We extensively tested and validated the stability of FPro in several databases such as the US Food and Nutrient Database for Dietary Studies (FNDDS) and the international Open Food Facts. FPro allowed us to implement an in-silico study based on US cross-sectional population data, where we showed that on average substituting only a single food item in a person’s diet with a minimally processed alternative from the same food category can significantly reduce the risk of developing metabolic syndrome (12.25% decrease in odds ratio) and increase vitamin blood levels (4.83% and 12.31% increase of vitamin B12 and vitamin C blood concentration) [37].

FoodProX takes as input 12 nutrients reported in the nutrition facts (Table S1), and returns FPro, a continuous score ranging between 0 (unprocessed foods like fruits and vegetables) and 1 (UPFs like instant soups and shelf-stable breads). We used the manual NOVA classification applied to the USDA Standard Reference (SR) and FNDDS databases to train FoodProX. In the original classification, NOVA labels were assigned by inspecting the ingredient list and the food description, but without taking into account nutrient content.

FPro does not assess individual nutrients in isolation but, rather, learns from the configurations of correlated nutrient changes within a fixed quantity of food (100 grams) [37]. Consequently, a single high or low nutrient value does not dictate a food’s FPro but the final score depends on the likelihood of observing the overall pattern of nutrient concentrations in unprocessed foods versus UPFs. For instance, while fortified foods may mirror mineral and vitamin content in unprocessed foods, our algorithm identifies unique concentration signatures unlikely to be found in minimally processed foods, resulting in a higher FPro [37].

The calculation of FPro for all foods in GroceryDB represents a generalization task, where the model faces “never-before-seen” data [71, 73]. More details on the training dataset, including class heterogeneity and imbalance, are available in Section S4.

### Price for calories trends

We applied robust linear models with Huber’s t-norm [74–76] to calculate regression coefficients and p-values for the relationship *log*(*PricePerCalorie*) ∼ *log*(*FPro*). The detailed regression results for each food category are presented in Figure S8, while the overall trend across GroceryDB is depicted in Figure 3A. To illustrate the price disparity at the extremes of food processing, the percentage change in price per calorie shown in Figure 3E was calculated by comparing the average price per calorie of the top 10% minimally processed items to that of the top 10% ultra-processed items within each category.

### Ingredient Trees

An ingredient list is a reflection of the recipe used to prepare a branded food item. The ingredient lists are sorted based on the amount of ingredients used in the preparation of an item as required by the FDA. An ingredient tree can be created in two ways: (a) with emphasis on capturing the main and sub-ingredients, similar to a recipe, as illustrated in Figure S16A; (b) with emphasis on the order of ingredients as a proxy for their amount in a final product, as illustrated in Figure S16B, where the distance from the root, *d*, reflects the amount of an individual ingredient relative to all ingredients. We opted for (b) to calculate IgFPro, as ranking the amount of an ingredient in a food is essential to quantify the contribution of individual ingredients to ultra-processing. In Eq. 1, we used 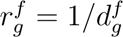 to rank the amount of an ingredient *g* in food *f*, where 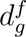 captures the distance from the root (Figure S16B for an example). Finally, IgFPro shows a remarkable variability when compared to the average FPro of products containing the selected ingredient (Figure S17), suggesting distinctive patterns of correlation between the products’ FPro and the ranking of ingredients in their ingredient lists.

### Database Structure

The database comprises two main files, both stored in CSV format for ease of use and accessibility:

1. GroceryDB Foods File This file contains comprehensive information about all the foods included in the GroceryDB. Each row represents a distinct food item. This file includes the following columns:
  - **name**: The name of the food item, typically as it appears on the product packaging.
  - **brand**: The brand or manufacturer of the food item.
  - **harmonized single category**: The general category or type of food (e.g., seafood, cereal, etc.).
  - **store**: The retail store where the food item is available (e.g., Walmart, Target, Whole Foods).
  - **f FPro**: Average FPro score of the food across the ensemble of classifiers. The FPro score is calculated using the FoodProX algorithm, taking into account the nutrition facts of the food.
  - **f FPro P**: a string indicating if the food has enough nutritional descriptors as detailed in SI Section 4.
  - **f min FPro**: Minimum FPro score across the ensemble of classifiers.
  - **f std FPro**: The standard deviation of the FPro score across the ensemble of classifiers.
  - **f FPro class**: expected NOVA class assigned according to FoodProX.
  - **ingredientList**: A list of ingredients used in the food item, providing insight into its composition and processing level. The ingredient list is crucial for calculating the IgFPro.
  - **has10 nuts**: boolean value indicating if the food is described by the 10 key nutrients described in SI Section 4.
  - **is Nuts Converted 100g**: Indicator if the food nutrients are converted per 100 grams.
  - **nutritional information**: Detailed nutritional information for the food item, including protein, total fat, carbohydrate, total sugars, total dietary fiber, calcium, iron, sodium, vitamin C, cholesterol, total saturated fatty acids, and total vitamin A.
  Please note that the prices of the food items are not included in this public release due to potential restrictions on public disclosure. However, we are willing to provide price information upon request. The file is available at https://github.com/ Barabasi-Lab/GroceryDB/blob/main/data/GroceryDB foods.csv.
2. GroceryDB IgFPro File This file contains data related to the IgFPro score of the ingredients listed in GroceryDB. Each rowcorresponds to a specific ingredient. The file is available at https://github.com/Barabasi-Lab/GroceryDB/blob/main/data/GroceryDB IgFPro.csv. The columns in this file are as follows:
  - **ingredient name**: The standardized name of the ingredient.
  - **count of products**: The total number of products in the database that contain this ingredient.
  - **ingredient FPro**: IgFPro calculated for the selected ingredient.
  - **average FPro of products**: The average FPro score of the products containing the selected ingredient.
  - **average distance to root**: The average distance of the ingredient from the root in the ingredient tree, representing its relative amount in the food item. Ingredients closer to the root contribute more significantly to the calculation of IgFPro.
  - **ingredient normalization term**: A numerical value used to normalize a food’s contribution to the IgFPro score, based on the ingredient’s overall ranking across all foods.

### Substitution Algorithm at TrueFood.Tech

TrueFood.Tech provides food substitution recommendations aimed at gently nudging consumers towards less processed alternatives. To accomplish this, we first identify food items that belong to the same category and share partial semantic similarity with the targeted item (range 0.10-0.95), based on both food names and ingredient lists. This approach increases the diversity of displayed recommendations while ensuring they remain within the same category.

We utilize the popular term frequency–inverse document frequency (Tf–idf) algorithm to measure the significance of words to foods in our database, adjusted for commonality across entries [77]. The similarity between weighted word vectors is calculated using cosine similarity. The final similarity between the queried food and other food items is determined by multiplying the ingredient-list-based similarity and the food-name-based similarity.

Next, we sort the semantically filtered foods by their FPro scores, ranking the recommendations in ascending order of FPro. This method allows us to identify the most similar food items with a lower FPro compared to the targeted item. Up to 50 items, listed in increasing order of FPro, are displayed on the website.

## Acknowledgments

We thank Dwijay Shanbhag at Northeastern University for his help on data collection and cleaning. We thank Daria Koshkina for help in designing the figures. A.-L.B is partially supported by NIH grant 1P01HL132825, American Heart Association grant 151708, and ERC grant 810115-DYNASET. G.M. is supported by NIH/NHLBI K25HL173665 and AHA 24MERIT1185447.

## Competing Interests

A.-L.B. is the founder of Scipher Medicine and Naring Health, companies that explore the use of network-based tools in health and food, and Datapolis, that focuses on urban data. All other authors declare no competing interests.

## Code and Data Availability

All code and data are available at BarabasiLab GitHub repository via https://github.com/Barabasi-Lab/GroceryDB/. Furthermore, GroceryDB is available to the public and consumers at http://TrueFood.Tech/.

## Author contributions

G.M., B.R., and A.-L.B. conceived and designed the research. B.R. performed data collection, data modeling, statistical analysis, data querying and integration, and contributed to the writing of the manuscript. G.I. and M.S. performed data cleaning, data curation, code cleaning and optimization, fact checking, and contributed to the writing of the manuscript. P.M. performed data cleaning, data integration, and contributed to the writing of the manuscript. G.M. and A.-L.B. wrote the manuscript and contributed to the conceptual and statistical design of the study.

## SUPPLEMENTARY INFORMATION

### 1 Motivation and Scientific Basis Behind the Development of FPro

The food matrix embodies a complex interplay of chemical components, influencing the release, transfer, accessibility, digestibility, and stability of numerous food compounds [1]. Ongoing research highlights how altered matrices in UPF may affect nutrient availability, postprandial glycemic response, and satiety levels [2–6]. The multifaceted impact of UPF on human health led us to explore concepts like metabolic response and food matrix through the lens of network science and ML, which account for critical dependencies and transcend reductionist single-nutrient analyses [7]. Indeed, by performing largescale analyses of nutrient concentrations in the food supply, we have documented how their amounts in unprocessed foods are constrained by physiological ranges determined by biochemistry and how different nutrient alteration patterns indicate reproducible food processing fingerprints [8, 9]. These findings have inspired and supported the development of FPro, which does not assess individual nutrients in isolation but, rather, learns from the configurations of correlated nutrient changes within a fixed quantity of food (100 grams) [10]. Consequently, a single high or low nutrient value does not dictate a food’s FPro but the final score depends on the likelihood of observing the overall pattern of nutrient concentrations in unprocessed foods versus UPF. For instance, while fortified foods may mirror mineral and vitamin content in unprocessed foods, the algorithm identifies unique concentration signatures unlikely to be found in minimally processed foods, resulting in a higher FPro [10].

### 2 Food Sources in NHANES

The majority of people rely on grocery stores as the primary source of food. To investigate this hypothesis, we looked into the National Health and Nutrition Examination Survey (NHANES), offering the variable DR1FS that corresponds to “Where did you get (this/most of the ingredients for this)?”, found at (https://www.n.cdc.gov/Nchs/Nhanes/2017-2018/DR1IFFJhtm#DR1FS). It is found that over 60% of all foods reported by NHANES 2017-2018 participants are from stores (Figure S1), indicating the high degree of reliance of the US population on grocery stores.

**Figure S1:**
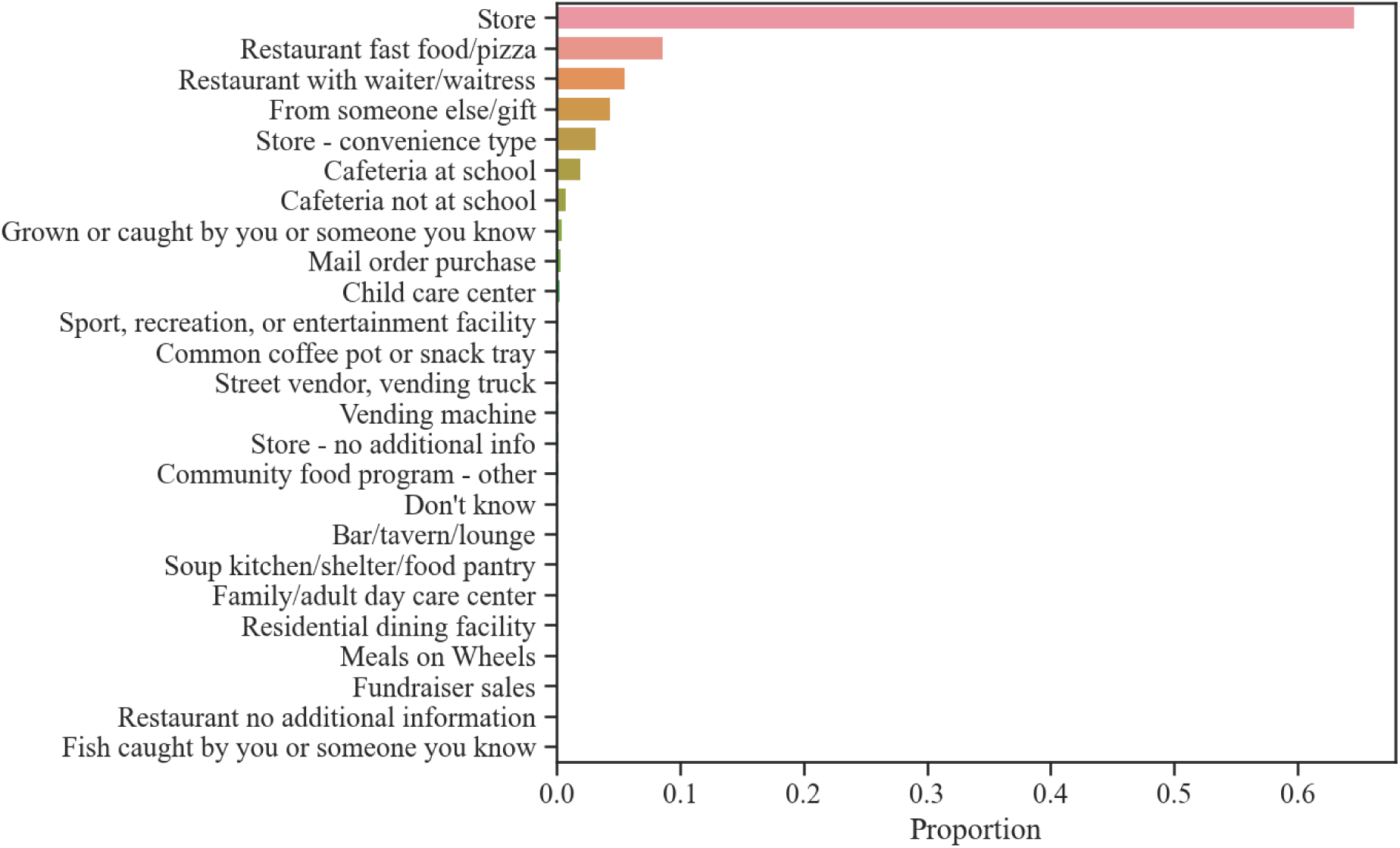
Proportion of Food Sources Reported in NHANES 2017-2018.

### 3 Data Collection and Processing

GroceryDB is built by collecting information regarding branded products from the publicly available data on the online websites of Walmart, Target, and Whole Foods. For data storage, MongoDB is used which offers highly flexible data structures with high input/output throughput.

#### 3.1 Identification of Additives

The “Substances Added to Food” database provided by the US Food and Drug Administration (FDA) is used as our primary dictionary to identify additives and their synonyms in food products [11]. In addition, the “Dictionary of Food Ingredients” (DFI) is used to further enrich and categorize the identification of additives [12]. Non-trivially, many substances declared on the nutrition fact labels have a broad range of synonyms. Thus, in addition to the synonyms provided by FDA, many synonyms are manually curated to better clean and normalize ingredient lists.

A major issue with cleaning ingredient lists is the high level of mismatch between labels provided by the FDA and the declared ingredients on nutrition fact labels printed by food producers [13]. This is aligned with the GS1 UK data crunch analysis, reporting 80% inconsistency in products data in the UK grocery industry [14]. This inconsistency could be partially explained by the high level of difficulty to find the common name of an additive. For example, the FDA food labeling guide encourages the use of common names, stating “always list the common or usual name for ingredients unless there is a regulation that provides for a different term. For instance, use the term ‘sugar’ instead of the scientific name ‘sucrose’ [15].” However, the FDA does not provide a strictly standardized database on the common names and synonyms of additives. While building GroceryDB, we frequently faced the issue that common ingredient names were not used on food packages. For example, the additive commonly known as “baking soda” is frequently declared as “sodium bicarbonate” on product labels. Similarly, “carmine”, a common coloring additive, is found in GroceryDB both as its standard name, “carmine”, and as “cochineal extract”, named after the insect at the origin of its red color. Lastly, as a note on terminology, the FDA distinguishes between “additives” and “substances added to food.” In this analysis we equate the label “additive” to the FDA’s “substance added to food.”

### 4 Food Processing Score (FPro)

FoodProX is used, a random forest classifier, to calculate FPro for the branded products in GroceryDB [10]. To train FoodProX, the manual NOVA classification on two USDA datasets is used, namely, FNDDS and Standard Reference (SR). For items with manual NOVA1, NOVA2, and NOVA3, we combined all unique nutrient profiles (12 nutrients) from FNDDS 2001-2018 (9 cycles), plus all the unique nutrient profiles from SR 20-28 (9 versions). This allows for more nutrient profiles among unprocessed and processed foods. However, for items with manual NOVA4, we only combined the latest nutrient profile for each food code from FNDDS 2001-2018 and SR 20-28. The reason for not including all unique nutrient profiles in NOVA4 class is to balance the training dataset, otherwise including all unique profiles in NOVA4 would make the training data from NOVA4 extremely dominant, not giving the classifier a chance to learn the characteristics of nutrient profiles in NOVA1-2-3 classes. The fraction of foods in each NOVA class is represented in Figure S2. The addition of SR database to the training data increased the number of training samples for NOVA1 and NOVA3 classes, hence balancing the training dataset.

The 12 nutrients in Table 1 is used to train FoodProX, since FDA requires reporting these nutrients on nutrition fact labels [16]. Although providing the minimum of 12 nutrients is mandated by the law, not all food labels declare those nutrients. Hence, we decided to rely on 10 nutrients and assume value 0 for Vitamins C and A, if these vitamins are not reported. The number of products that reported at least 10 nutrients in GroceryDB is shown in Figure 3. For consistency, we decided to ignore all foods that do not meet the minimal requirement of 10 nutrients [17]. If necessary, the value of missing nutrients could be imputed to measure the degree of food processing for all foods currently excluded.

**Figure S2:**
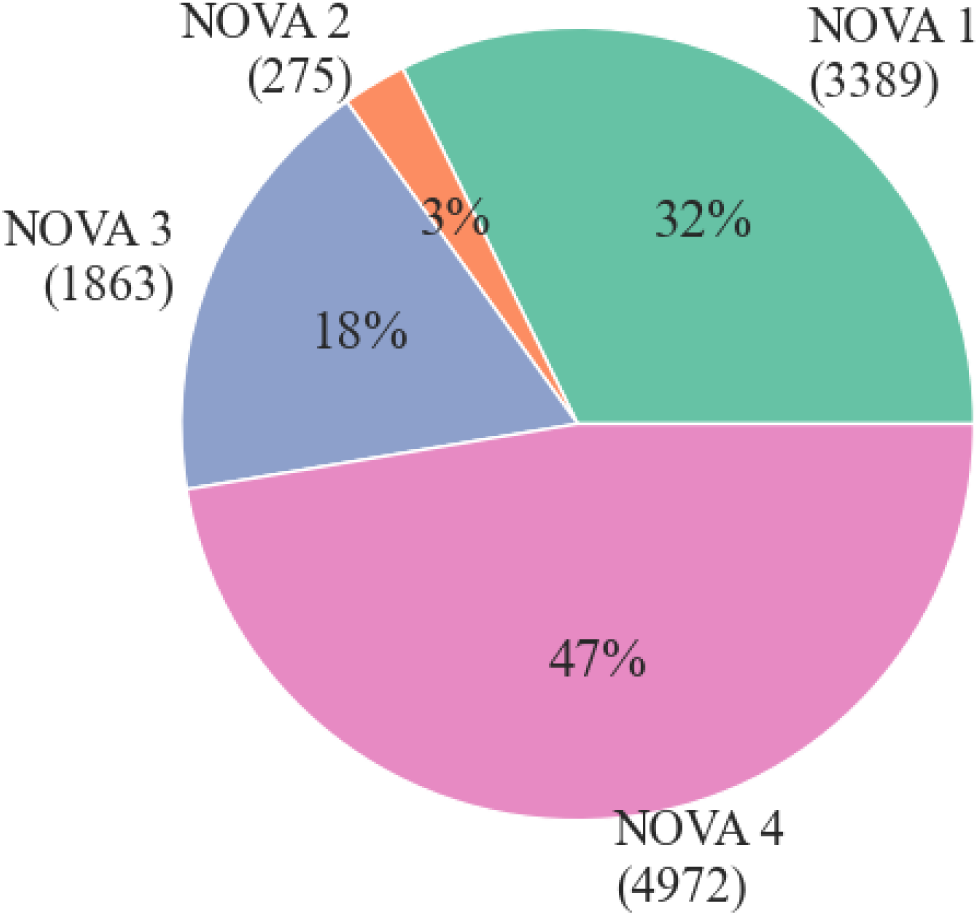
Fraction of NOVA Classes in the Training Dataset. NOVA classifications are used to train FoodProX [10], the ML classifier that assigns a FPro score to each food.

### 5 Comparison with Categorical Food Classification Systems

The current standard food processing classification systems are categorical systems that divide foods into multiple descriptive categories, commonly ranging between 4 to 8 categories that are manually designed to capture variability from unprocessed to ultraprocessed foods [18, 19]. Such categorical classification systems lead to considering many foods as equally ultra-processed, facing the conclusions that 60% of the global, 73% of the USA, and 80% of the South Africa food supply is ultra-processed [10, 20–22]. Recently, this has also led to 70.2% of all Greek branded food products being classified as UPF, with subcategories of foods included in the sustainable and traditional Mediterranean diet scoring 58.7% or 41.0%, respectively [23]. Furthermore, this homogeneity in food classification systems makes it difficult to address the impact of ultra-processing with substitution or reformulation strategies [24–27]. To address this issue, the SIGA classification developed a holistic-reductionist approach, which intends to offer a practical tool for the food industry [28]. SIGA defines an exhaustive list of markers of ultra-processing, helping to reduce the ambiguity in the interpretation of descriptive food labels. By utilizing these markers, SIGA expands the number of food processing categories to nine, introducing subgroups based on the degree of processing of ingredients, added salt and sugar, and fat contents. Despite these commendable efforts, SIGA still misses extensive epidemiological assessments over dietary intake surveys, and as a categorical classification system leaves open questions regarding the appropriate number and types of processing categories and the amount of information necessary to reliably identify them. A continuous food processing score eliminates the need for identifying categories of food processing by capturing maximal variability in the degrees of food processing. Such variability enabled us to derive the distributions of food processing scores across multiple grocery stores (Figure 2) and observe that, in soups & stews, a 10% increase in FPro results in 24.3% decrease in price per calorie. Furthermore, we were able to 1) test the stability of FPro with varying selections of nutrients, 2) validate its robustness against the expected variability and uncertainty in nutrient content, and 3) gain higher predictive power in epidemiological studies compared to categorical classifications, uncovering associations [10].

**Table S1:**
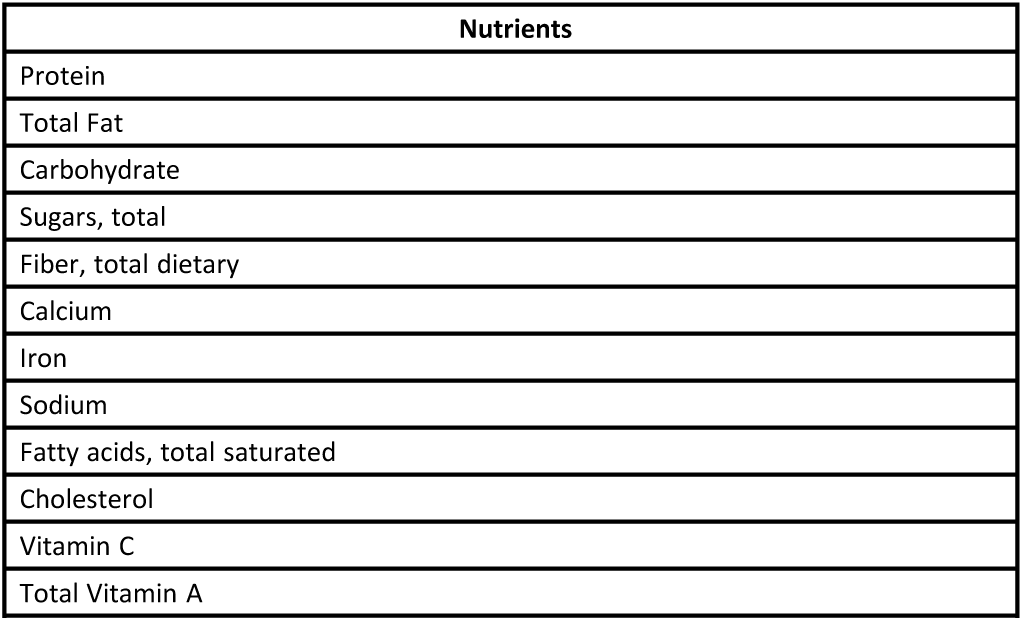
12 Nutrient Panel for Branded Products.

| Nutrients |
| --- |
| Protein |
| Total Fat |
| Carbohydrate |
| Sugars, total |
| Fiber, total dietary |
| Calcium |
| Iron |
| Sodium |
| Fatty acids, total saturated |
| Cholesterol |
| Vitamin C |
| Total Vitamin A |

**Figure S3:**
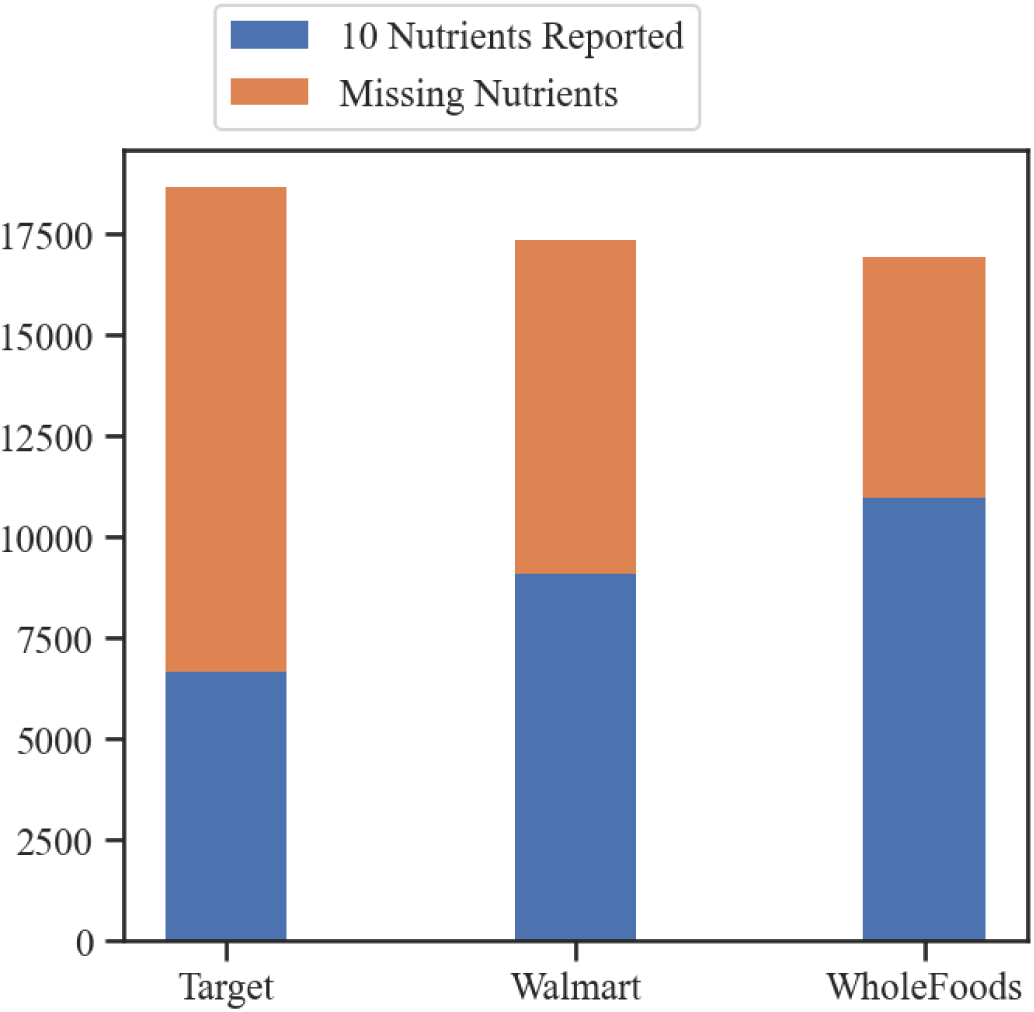
Number of Products with Missing Nutrients in GroceryDB. The products that did not report one of the following 10 nutrients are marked as ‘missing nutrient’: protein, total fat, carbohydrate, total sugars, total dietary fiber, calcium, iron, sodium, total saturated fatty acids, and cholesterol.

### 6 Comparison with USDA FoodData Central and Open Food Facts Databases

GroceryDB is matched with the USDA FoodData Central Global Branded Food Products Database (BFPD) according to the product name and the ingredient lists. We find that BFPD only covered 44% of the products in GroceryDB with a *Similarity Score* ≥ 0.95 (Figure S4A). We performed that same analysis with OpenFoodFacts (OFF), finding OFF only covers 38% of GroceryDB (Figure S4B).

Interestingly, the products missing components of the nutrient facts in GroceryDB are also missing the data within BFPD. Specifically, within the 22,900 items with missing nutrition facts in BFPD, only 9,600 were matched with *Similarity Score* ≥ 0.95 to GroceryDB. From these 537 had full nutrition facts within GroceryDB. These findings suggest that GroceryDB offers a more up to date picture of the food supply than BFPD, and many products do not report the full nutrient panel required by the FDA.

**Figure S4:**
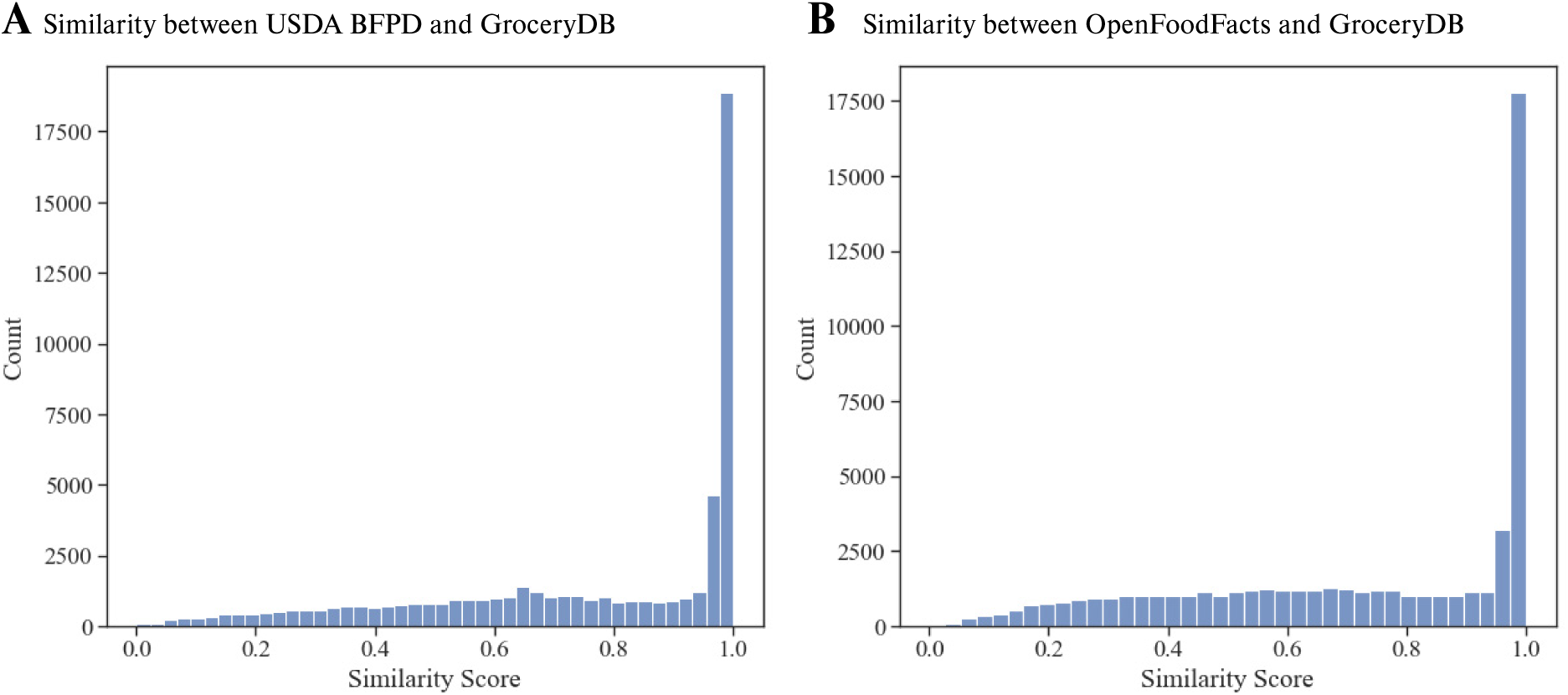
Coverage Comparison with USDA BFPD and OpenFoodFacts. GroceryDB offers a complementary picture of the food supply compared to BFPD and OFF. **(a)** Similarity scores are based on ingredient lists declared in BFPD and GroceryDB, using the TF-IDF (Term Frequency-Inverse Document Frequency) algorithm (see Methods, Substitution Algorithm at TrueFood.Tech). The BFPD (April 2021 version) has 1,142,610 branded products and GroceryDB has 50,467 items from which 2,754 items are excluded because of not having an ingredient list. The similarity score is calculated for the remaining 47,713 items, finding that BFPD only covers 44% of the products in GroceryDB with *Similarity Score* ≥ 0.95. **(B)** Similarly, the overlap between OFF and GroceryDB is investigated. Among 426,479 products in OFF with English list of ingredients (as of January 2022), only 18,948 products exist in the entire GroceryDB with *Similarity Score* ≥ 0.95, covering 38% of the products in GroceryDB.

### 7 Category Harmonization

Grocery stores classify foods into multiple categories and sub-categories, for a total of over 200 main categories and 866 sub-categories that are hierarchically organized in levels. Grocery store categories tend to be organized according to the store layout, helping consumers navigate the store. In contrast, epidemiological databases tend to categorize foods based on processing methods and the origin of food (type of plant or animal parts). For instance, FNDDS 2017-2018 has multiple categories for milk: ‘Milk and Milk Products’, ‘Milks, milk drinks, yogurts, infant formulas’, ‘Milk, fluid, evaporated and condensed’, and ‘Milk, fluid, imitation’ (declared by first 5 digits of food codes). Another approach to food classification is used by the What We Eat in America (WWEIA) database, aiming to provide categories that better resonate with consumers. For example, WWEIA contains 10 categories for milks, separating milk flavors and fat concentrations, ranging from ‘Milk, whole’, ‘Milk, reduced fat’, and ‘Milk substitutes’, to ‘Flavored milk, whole’, ‘Flavored milk, nonfat’, and ‘Milk shakes and other dairy drinks’ [29]. In GroceryDB, a similar approach as WWEIA is followed, with additional emphasis on the consumer’s use of products to enable effective food substitution strategies. For instance, meat-based and plant-based burgers are placed into a single category, ‘Prepared Meals & Dishes’, since from the consumer perspective these are both ready-to-cook burgers. This method of categorization leads to broader food categories and higher food variability, allowing more opportunities for meaningful food substitution recommendations.

The foods from grocery stores are harmonized into 42 broad categories designed for assisting food recommendation algorithms that aim at finding alternative food choices within the same category. For instance, in grocery stores the *frozen-foods* category includes items ranging from frozen fruits and vegetables to frozen lasagna and breakfast egg bites. Therefore, we broke the *frozen-foods* category into “Packaged Produce”, “Breakfast”, and “Prepared Meals & Dishes”. The list and size of the harmonized categories in GroceryDB are shown in Figure S5. The reason for observing such a large number of categories in stores is due to the lack of a standard classifying method across the stores. For example, breads as Level 1 category are marked as “bread bakery”, “bakery bread”, and “breads rolls bakery” in Walmart, Target, and Whole Foods, respectively and only diversify more by sub-category (Figure S6).

Lastly, since the focus of this paper is investigating the extent of food processing in grocery stores, we decided to not include the categories that are naturally unprocessed in all analysis, except in Figure 2A and Figure 2C that illustrate the distributions of FPro. Food categories such as fresh produce, raw beans, eggs, and raw meat are examples of categories that are naturally unprocessed.

### 8 Price and Food Processing

To investigate the hypothesis that processing impacts food prices, the price per calorie of the branded products are calculated as the total package price divided by the package calories (Figure S7A-B). Items with zero calories like Coke Zero are ignored in this analysis. Next, the Spearman’s correlation coefficient between FPro and price per calorie is calculated, as captured by the correlation matrix in Figure S7C. Depending on the store and categories, the food processing correlates with cheaper calories, as in the case of the strong negative correlation for breakfast, mac-cheese, pudding-jello, cakes, and pastrychocolate-candy. Finally, in a few categories like milk-milk-substitute, pasta-noodles, cheese, and jerky the more processed foods tend to have a higher price (Figure S7C).

**Figure S5:**
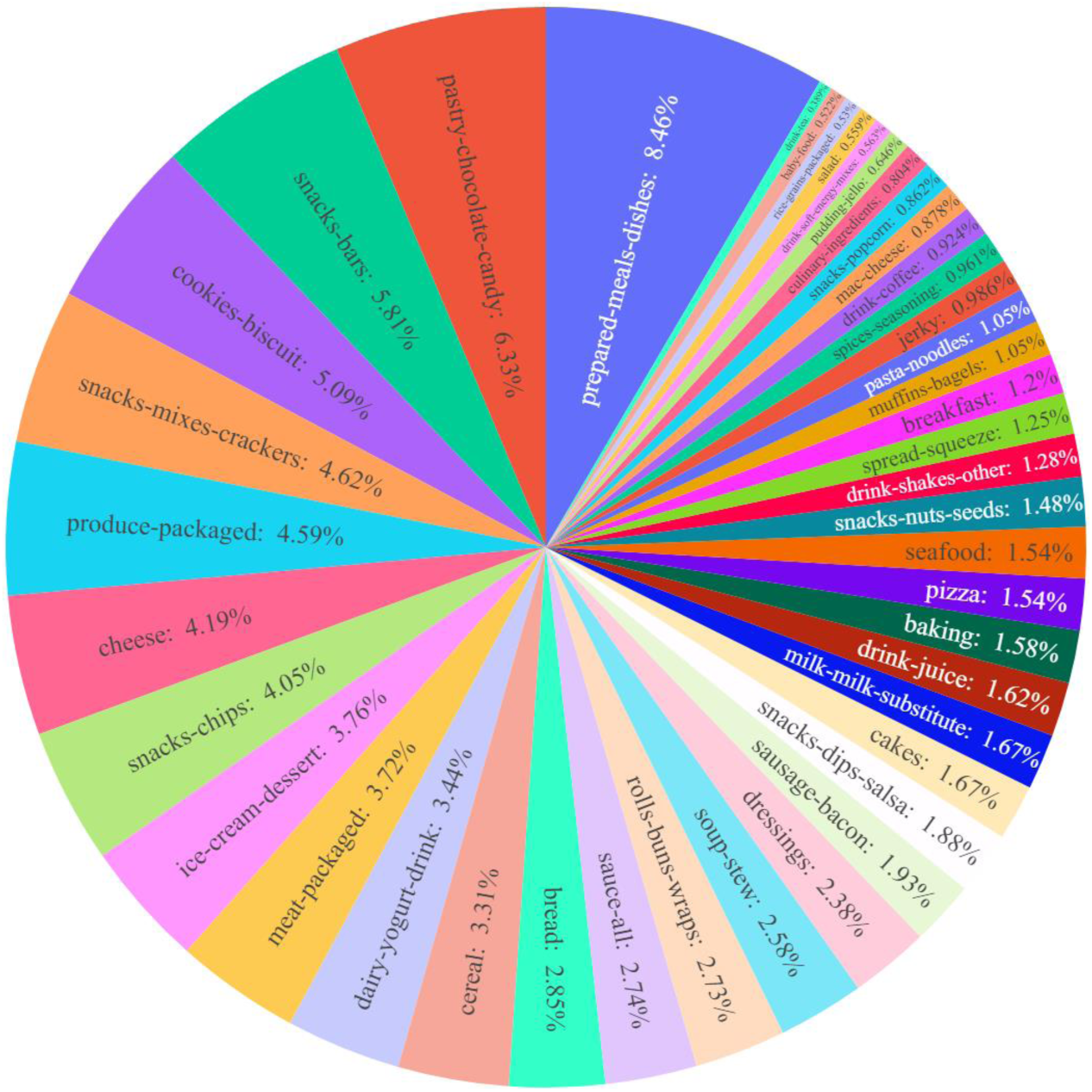
Fraction of Foods in Harmonized Categories. We harmonized foods from the three grocery stores into coarse-grained categories to represent foods from the perspective of individuals searching for alternative food choices. A comparison between store categories and GroceryDB categories is illustrated in Figure S6.

To further assess the relationship between FPro and price per calorie, robust linear models are used [30, 31]. The regression coefficients and p-values are illustrated in Figure S8. Note that Figure 3 does not include the pasta-noodles category. While this category contains enough data points to perform regression analysis, it does not exhibit a Gaussian-like distribution for residuals. Furthermore, the FPro values for pasta-noodles are highly segmented, not covering a broad range of FPro bins as seen in the other categories (Figure S9). Considering the limitations of the available data, it is found that on average the highly processed pasta-noodles are 46% more expensive than minimally processed-alternatives (comparing the averages in the top and bottom 10% of FPro), capturing a trend that goes from raw uncooked pasta to instant noodles, fortified pasta, and seasoned pasta.

**Figure S6:**
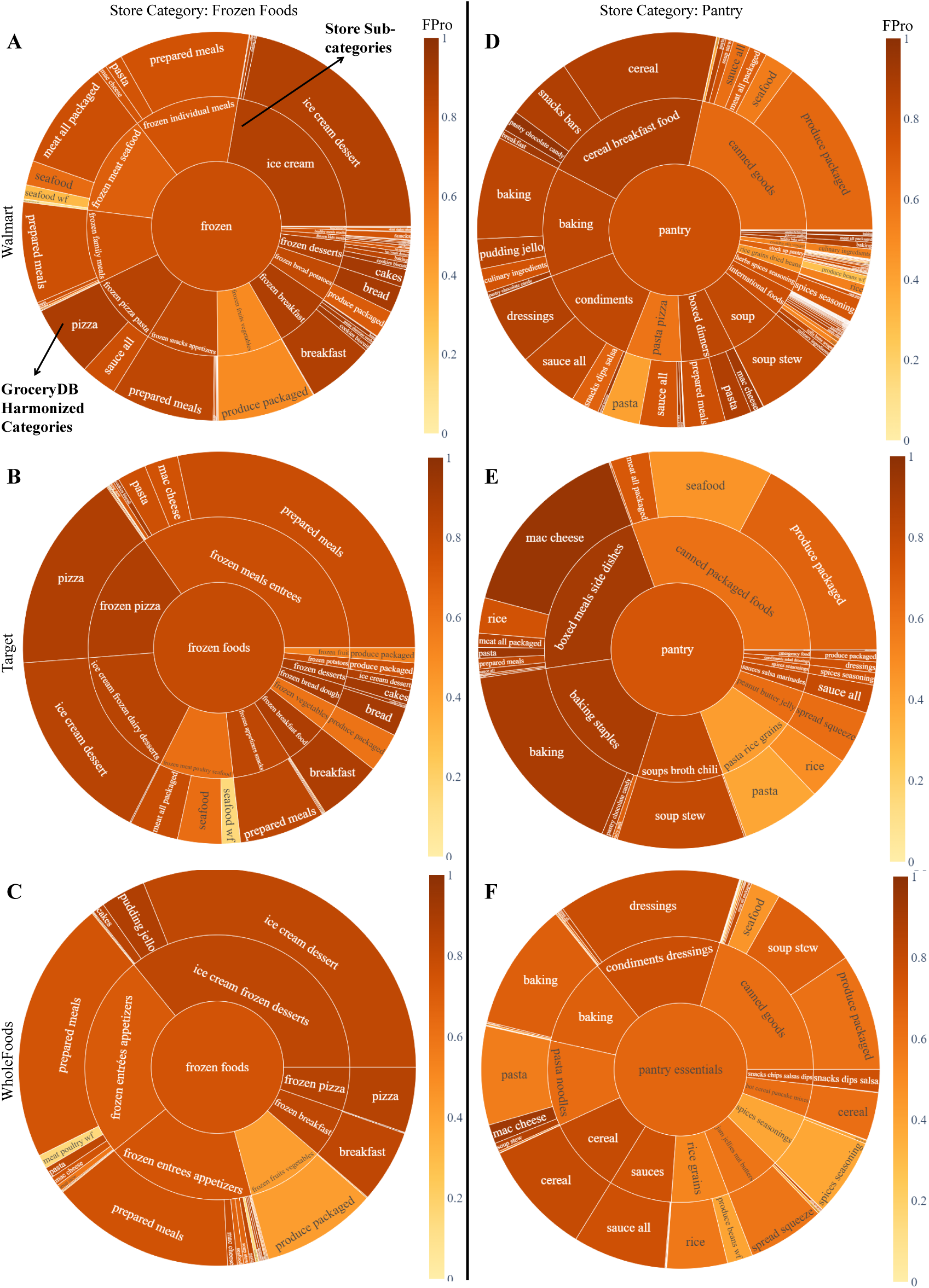
Store Categories vs Harmonized Categories. **(A-F)** Breakdown of frozen and pantry foods commonly found in the category hierarchy of grocery stores (inner cycles), mapped onto the GroceryDB harmonized categories (outer cycle). Colorbar shows the degree of processing of each category using FPro.

**Figure S7:**
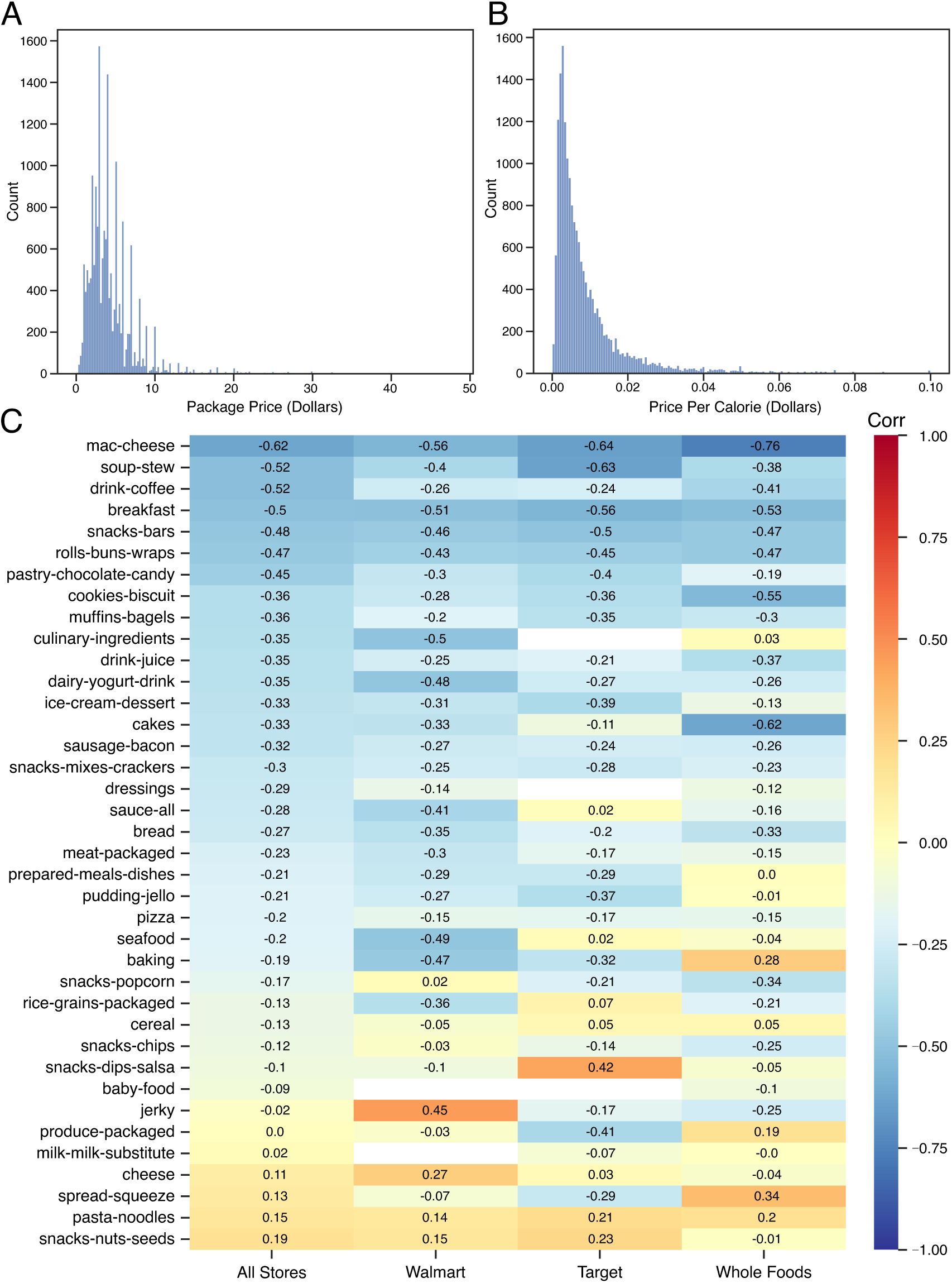
Correlation between Price and Degree of Food Processing. **(a)** The distribution of price per item in GroceryDB. **(b)** The distribution of price per calorie obtained by dividing an item’s price by its total calories (zero calories items are not included in this analysis). **(c)** The Spearman’s rank correlation coefficient between price per calorie and FPro across food items. An higher extent of food processing tends to decrease cost in many categories, but not in all of them.

**Figure S8:**
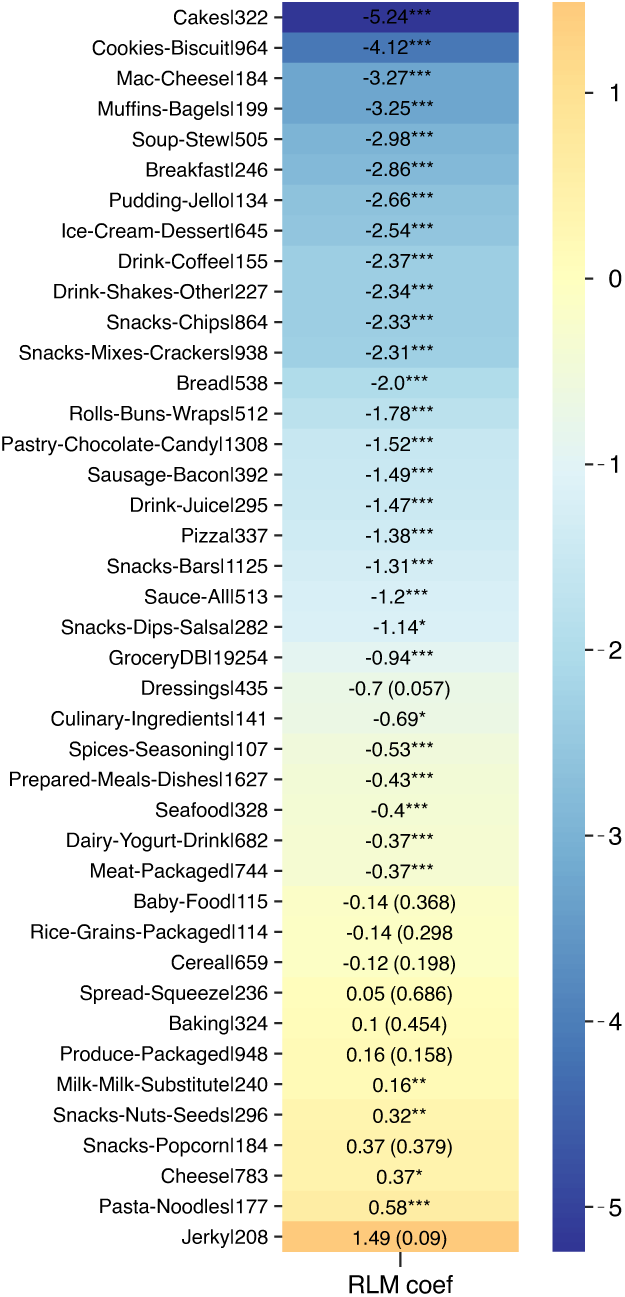
Price and Degree of Processing. Regression coefficients and p-values for *log*(*PricePerCalorie*) ∼ *log*(*FPro*) using robust linear models, a two-sided test, to assess the relationship between price and FPro in GroceryDB [30,31] (* = p-value ≤ 0.05, ** = p-value ≤ 0.01, *** = p-value ≤ 0.001). The p-values for non-significant results are shown in parentheses. The number to the right of each category represents the number of foods contributing to the regression, having metadata for package weight, total calories, and nutrients necessary to calculate FPro. No adjustments were made for multiple comparison given the small size of the dataset.

### 9 Organization of Ingredients

This section describes the methods developed to quantify the organization of ingredients in the food supply.

**Figure S9:**
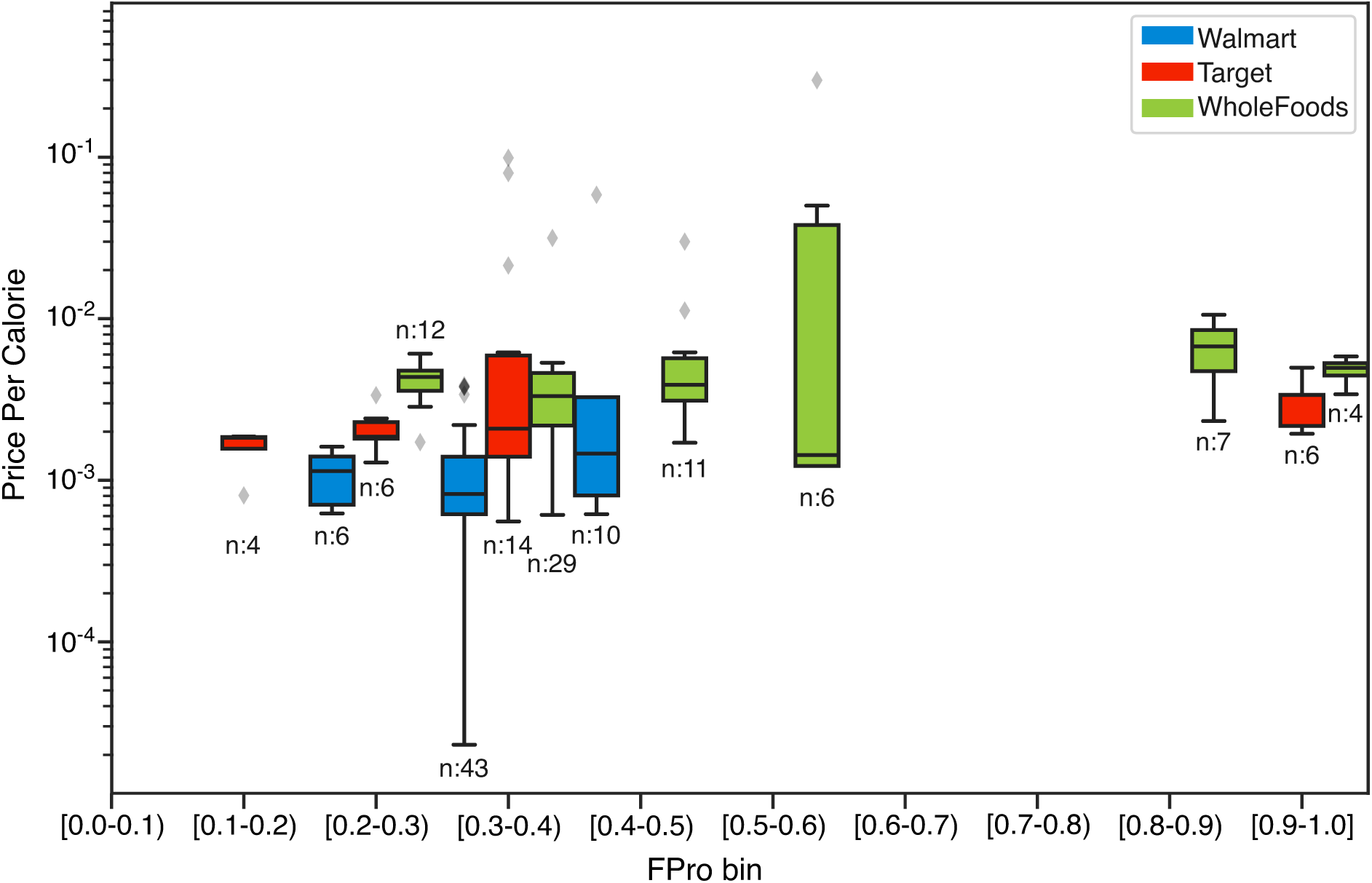
FPro vs. Price Per Calorie for Pasta & Noodles Category. In the pasta-noodles category, only 38.6% have sufficient metadata to estimate the relationship between FPro and price per calorie. While the number of data points is enough to perform a regression analysis, this category does not exhibit a Gaussian-like distribution for residuals. Furthermore, the FPro values for pasta-noodles are highly segmented, not covering a broad range of FPro bins as seen in the other categories. For the box plots, the minimum is the lower quartile, the central line is the median, and the maximum is the upper quartile. The whiskers show data outside of the inter-quartile range. Diamonds represent outliers.

#### 9.1 Cleaning Ingredient Lists

The ingredient lists are regulated by the FDA food labeling guidelines, however there are numerous nuances that cause inconsistency and require normalization [32]. For example, corn starch is reported as “cornstarch”, “corn-starch”, “corn starch”, and “starch made from corn” in the ingredient lists, and needs to be normalized to one format. Similarly, the FDA allows using a variety of synonyms for an ingredient, like the common synonyms “soybean”, “soy”, and “soya” used for soybeans. Ingredient lists often provide a variety of information for individual ingredients such as 1) processes involving in their preparation, e.g., “expeller-pressed canola oil”, where “expeller-pressed” describes how the canola oil is made, 2) intended purpose of their use, e.g., “calcium carbonate as a firming agent”, where “firming agent” describes why the compound was added to the product, and 3) alternative names, e.g., “vitamin b3 (niacinamide)”, where niacinamide is the chemical name for vitamin b3. Additionally, the FDA does not limit products to true ingredients, allowing products to have ingredients such as “smiles” and “pure love”. The heterogeneity and variability of declared labels in the ingredients lists requires a base-knowledge to organize the list of ingredients into ingredient trees.

We leverage the dictionary of food ingredients [12] and manually investigate the ingredient list of products in GroceryDB to curate a list of ingredients within the products. From this we found that the ingredient lists contain many miss spellings of the ingredients as well as syntax structural errors, such as neglecting a comma between two ingredients. We performed an bigram search between the ingredients to find and correct potential errors, generating a dictionary of changes to apply. We added ingredient synonyms to the dictionary to unify the names of each ingredient. This curation dictionary replaces strings within each ingredient list (Figure S10).

Generating ingredient trees from the reported ingredient lists will only work for products which respect the structural syntax where ingredients are ranked by their contributions to the final product. Products such as snack packs or variety packs contain several ingredient lists and thus there are multiple ingredient trees, making unclear the total contribution of each ingredient. In addition, this also applies to products with multiple components such as frozen pizzas where the topping, sauce, and crust offer individual ingredient lists. We remove these products from our analysis. Secondly, many products contain uneven brackets, meaning that where the main ingredient list and sub-ingredient list begins and ends is unclear. We compare these products to others with similar ingredient lists in order to correct the uneven brackets. Lastly, the separator between ingredients can vary between products and differ in sub-ingredient lists, therefore, we count the special characters in the lists and denote the most common character as the separator to use in splitting.

Conjunction words within the ingredient lists require specialized rules for splitting the list into individual ingredients. When “and” is present, we consider the rank of the ingredients to be as written; thus we replace “and” with a separator. If “or” or “and/or” is present then the ingredient list displays a list of options or possible combinations, not an exact list of ingredients. For example, “vegetable oil (canola oil and/or palm oil)” indicates that the vegetable oil can be either 1) canola oil, 2) palm oil, or 3) canola oil and palm oil. We capture this vagueness within our ingredient trees by applying the same rank to both canola oil and palm oil instead of giving palm oil a lower rank than canola oil.

**Figure S10:**
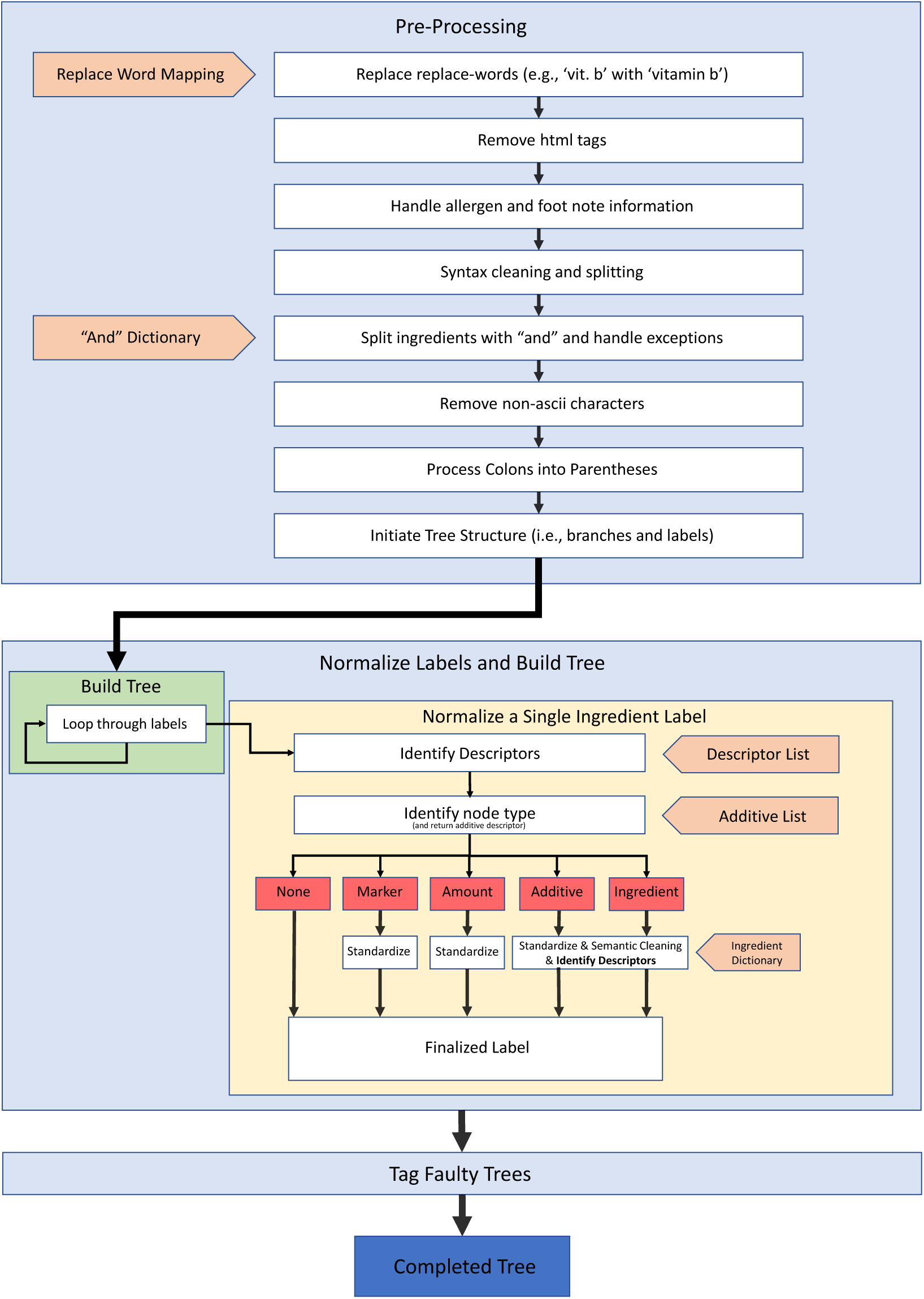
Pipeline for Transforming an Ingredient List into an Ingredient Tree. An extensive data engineering and cleaning is needed to harmonize the list of ingredients declared on foods in grocery stores. The lack of a standardized ontology of ingredients makes this task more difficult.

The descriptors of ingredients are captured by using part-of-speech tagging followed by manual inspection to correct errors since such algorithms are trained on sentences and not short phrases as well as are not fine-tuned on food specific terminology. We annotate descriptors of processing such as “expeller-pressed”, “hydrogenated”, and “partially hydrogenated” and this extra information is organized and annotated as “

*<*descriptor*>*” in the cleaned ingredient labels. Descriptors that do not indicate processing are removed since they would add unnecessary divisions of ingredients or layer to the ingredient tree. For example, in “vitamin b3 niacinamide”, the descriptor “niacinamide” would separate it from “vitamin b3”, however, niacinamide is just the chemical name of vitamin b3, thus we prevent separating these as distinct ingredients. Furthermore, “sodium acsorbate (acidity regulator)” places the purpose of the compound in brackets and since brackets indicate sub-ingredients, this would place the descriptor “acidity regulator” as a sub-ingredient. We remove these instances to prevent the unnecessary growth of ingredient trees.

The raw format of ingredients as reported on product packages contains over 32,000 unique labels, including a broad range of heterogeneous synonyms and semantically duplicate labels. Through our curation, we were able to reduce this number to approximately 19,600 labels corresponding to about 14,400 unique ingredients.

#### 9.2 Approximating the Number of Ingredients in GroceryDB

Given the high level of data inconsistency in the grocery industry [14], it is difficult to find the exact number of ingredients and additives in GroceryDB. Hence, we use the population proportion estimation statistics (Cochran’s sample size formula) to estimate the number of ingredients, additives, and descriptors. The number of unique ingredients is estimated to be 18,146 (Figure S11A), including 1,456 additives (Figure S11B). Two quantities are presented, one considering descriptors and another without descriptors, providing two levels of ingredient specificity. With descriptors, for example, the string “bleached wheat flour” is counted separately from “enriched wheat flour.” Without descriptors, both are considered “wheat flour.” Counts become larger when considering descriptors. Through this analysis we found additives like “corn starch” which appears in GroceryDB with 13 different unique descriptors like “modified,” and “resistant.”

The FDA identifies 3,972 total substances added to food along with potential synonyms [11], 1,316 of which are classified as direct, approved additives [33]. The Dictionary of Food Ingredients (DFI), an industry standard encyclopedia of food ingredients in the U.S. [12], lists 609 additives. The DFI is based on the FDA’s Title 21 in the Code of Federal Regulations [33], and there is thus a considerable overlap between the two sources. With the current pipeline and duplicate removal, DFI adds 205 additives beyond the FDA source. We use both DFI and the FDA’s database of additives to identify additive ingredients in GroceryDB, resulting in an estimated 843 unique additives not considering descriptors.

This task is difficult as there is not a one-to-one mapping between unique strings in the ingredient lists, and an actual ingredient. For example, for the ingredient “vitamin b12”, typos like “vitemin b12”, branded strings (“Walmart vitamin b12”), and alternate spellings (“vit. b12”) are found, which all point to the same ingredient. While our cleaning caught many of these issues, a full cleaning of ingredient strings remains a goal of future work, but an approximate number of unique ingredients in the U.S. food system with the current state of GroceryDB is achievable.

To estimate the number of ingredients and additives, samples of ingredient labels are collected after running the data cleaning pipeline (Figure S10) and selected ingredients present in at least two products. Then, manual investigations of these samples are done to count the number of labels that are correctly classified vs. incorrectly classified (due to the lack of a comprehensive dictionary of synonyms and the large level of data inconsistency in the grocery industry). Next, Cochran’s formula is used to estimate the number of ingredients and additives based on the proportion of correctly classified labels.

Finally, GroceryDB paves the way towards systematically quantifying the organization of ingredients in the food supply. Indeed, the level of data inconsistency is estimated to be 80% in the grocery industry [13, 14]. Given this high level of data inconsistency, future work will extend the current efforts on data cleaning, integration, and normalization, to better approximate the number of unique ingredients and additives in the food supply. Yet, GroceryDB provides the data structure, methods, and pipeline needed to systematically unify the ingredient lists in the food supply, and unveil the organization of ingredients.

**Figure S11:**
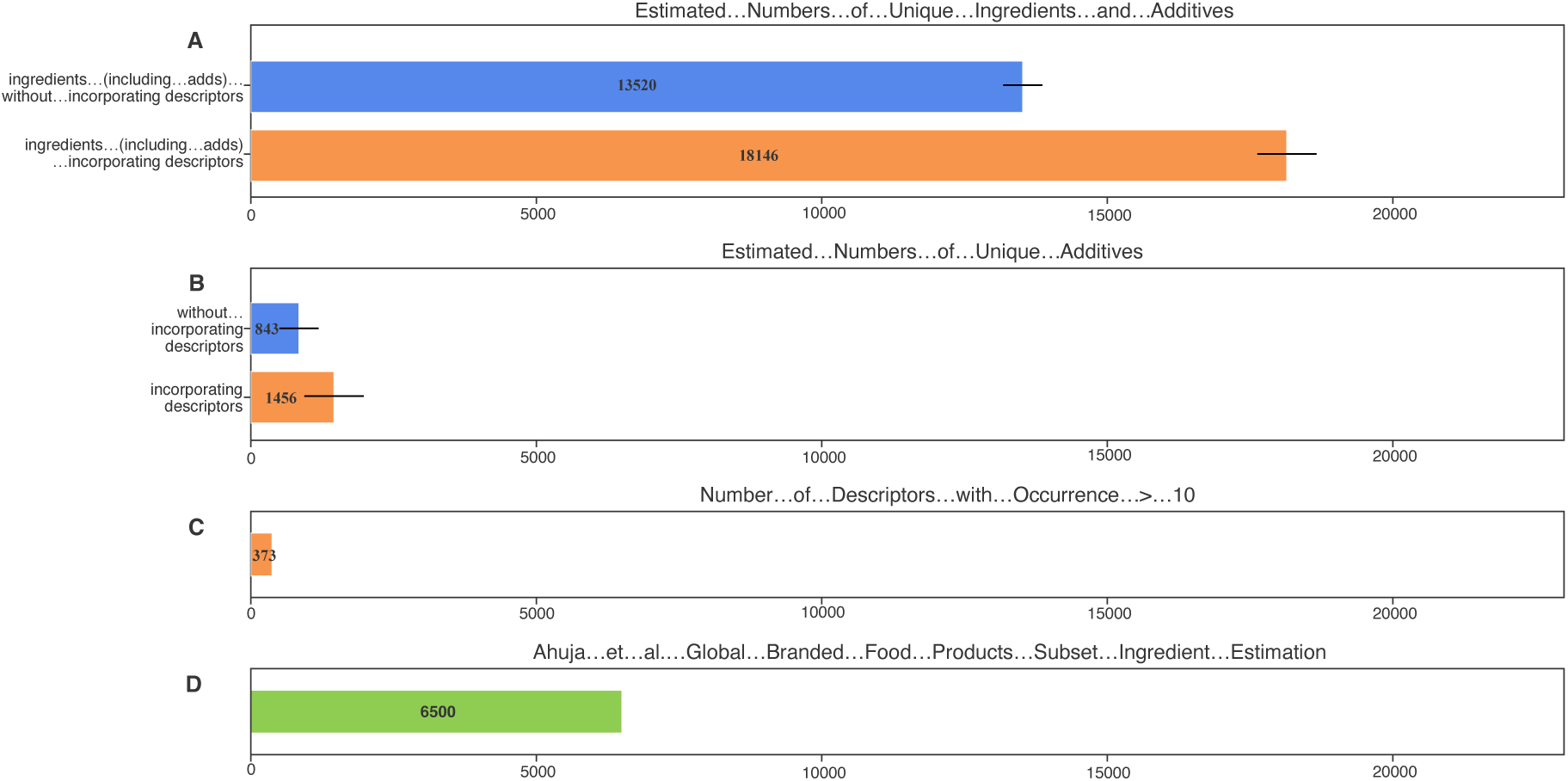
Estimated Number of Ingredients, Additives, and Descriptors in GroceryDB. **(A-B)** Estimation for the unique number of ingredients and additives in GroceryDB based on Cochran’s formula using random sampling of n = 377 ingredients from the total list of ingredients. The error bars are the 95% confidence interval of the Cochran’s formula. The descriptors add a significant complexity in estimating the number of ingredients and additives in the food supply. Examples of ingredients with descriptors are “bleached wheat flour” and “enriched wheat flour” where ‘bleached’ and ‘enriched’ are descriptors. **(C)** The number of descriptors that appeared at least in 10 products. Although we manually created a dictionary of descriptors with 168 labels, natural language processing is mainly relied on to automatically identify descriptors. This process resulted in identifying more descriptors. **(D)** Ahuja et al in [13] analyzed a subset of BFPD, resulting in the identification of 6,500 ingredients from 5 out of 31 food categories in BFPD. The data cleaning led to the identification of a smaller number of ingredients without descriptors, for a total of 4,201 ingredients.

#### 9.3 Characteristics of Ingredient Trees

The branded products with more complex list of ingredients are more likely to be highly processed. To test this hypothesis, two measures are found to characterize the ingredient trees: tree width and depth. The tree width, denoted by *W*, represents the number of main ingredients in a product, and the tree depth, *D*, approximates the extent of the reliance on mixtures of sub-ingredients. We define depth-sum of an ingredient tree, denoted by *D_s_*, as the sum of the depth of all its branches. Figure 5 presents two cheesecakes with *FPro* = 0.953 and *FPro* = 0.720 along with their corresponding ingredient trees. The highly processed cheesecake has a complex ingredient tree with *W* = 31 and *D_s_* = 3 (Figure 5A). In contrast, the less processed cheesecake has a simpler ingredient tree with *W* = 5 and *D_s_* = 2 (Figure 5B).

Theoretically, by the definition of branch depth sum, *D_s_* may be considered as another representation of *W*, if most main ingredients of products have sub-ingredients. Yet, *W* and *D_s_* show varying characteristics, depending on food categories. For example, cheeses tend to have high *W* and low *D_s_*, whereas cakes have relatively lower *W* and higher *D_s_*, showing distinct behaviors as illustrated in Figure S12A. In contrast, when comparing cakes and pizzas (Figure S12B), the difference between *W* and *D_s_* is less striking. Moreover, the distributions of *W* and *D_s_* show more differences than similarities as illustrated in Figure S13A-B, indicating that they are presenting different information. Interestingly, Whole Foods tends to offer foods with both smaller *W* and shorter *D_s_* compared to the other stores. That is, products in Whole Foods tend to have less ingredients and also rely less on mixing sub-ingredients.

Finally, we analyzed the relationship between *W* and *D_s_* of ingredient trees in all categories. The Spearman’s correlation between *W* and *D_s_* suggests that these metrics capture unique information about the products. There are both positive and negative correlations between *W* and *D_s_*, depending on the categories (Figure S13C). The strongest correlation between *W* and *D_s_* is in the culinary ingredients and spices-seasoning, since these are the categories that in principle should least rely on the mixture of sub-ingredients, as generally spices and culinary ingredients are made of simple ingredients. In milk & milk-substitutes, there is a strong correlation between *W* and *D_S_*, partially explained by the transition from simple whole milk to chocolate and milk-substitutes (like almond and oat milks), increasing the number of main ingredients and sub-ingredients (Figure S13). On the other hand, in breads, ice cream, and sausage, there is a negative correlation between *W* and *D_s_* partially explained by the use of more complex ingredients. For instance, flour, dough conditioner, and cookie dough are often reported with a long mixture of sub-ingredients, resulting in a higher *D_s_* in breads and ice creams.

**Figure S12:**
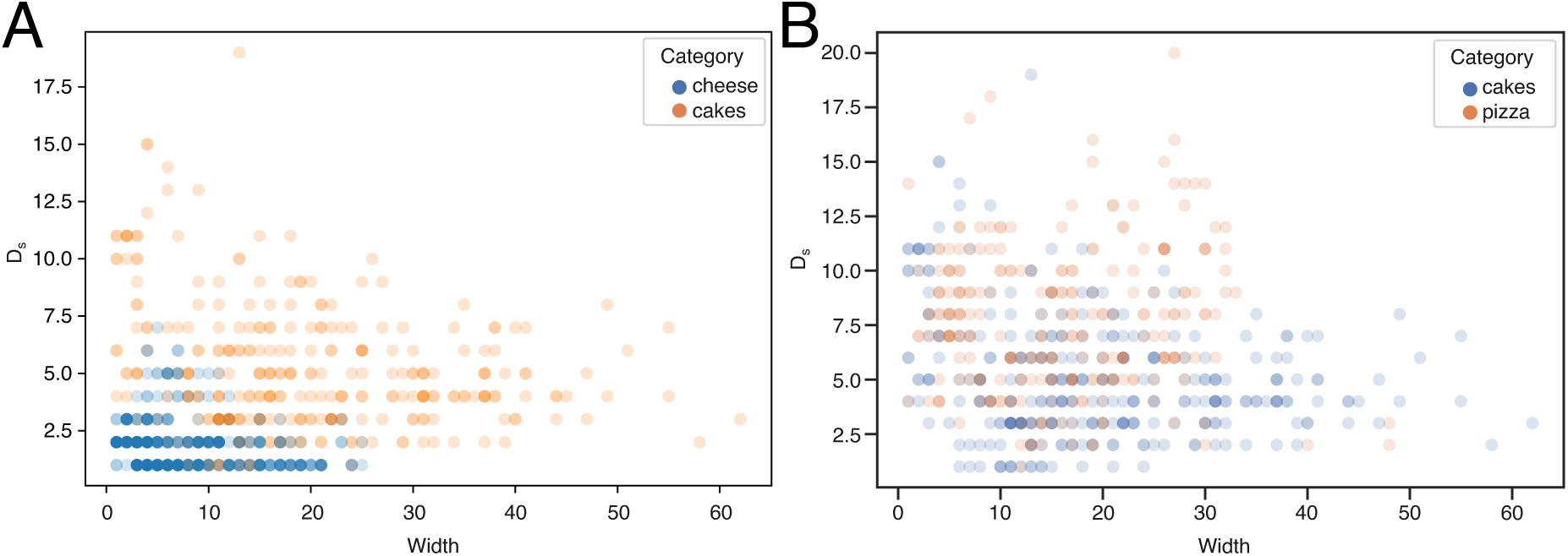
Tree Width vs Depth-Sum (*D_s_*). The ingredient trees behave differently according to food categories. **(a)** In the cheese category the ingredient lists tend to rely less on the mixture of sub-ingredients (wider trees), whereas in cakes we observe that more sub-ingredients are defined. **(b)** The ingredient trees of cakes and pizzas show similar structures.

#### 9.4 Correlation between Characteristics of Ingredient Lists and FPro

To test the hypothesis that branded products with more complex list of ingredients are more likely to be highly processed, we also investigate the relationship between *W*, *D_s_*, and FPro. For example, in cereals, pasta-noodles, and baking categories, the items that have simpler ingredient trees also have a significantly lower FPro (Figure S14A-C). However, this effect is weaker in prepared Meals & dishes, where lower values of FPro with relatively large ingredient trees are found (Figure S14D).

Lastly, the Spearman’s correlation between *W*, *D_s_*, and FPro is investigated. Generally, there is a strong correlation between *W*, *D_s_*, and FPro indicating that products with complex ingredient trees tend to have a higher FPro (Figure S15). Also, some categories show stronger correlation with *D_s_* and FPro, signaling that mixing many sub-ingredients may drive food processing. For example, in breakfast products, pizzas, popcorn, we find that *D_s_* has a stronger correlation with FPro compared to *W* and even the total number of ingredients (defined as the sum of all ingredients and sub-ingredients including additives, Figure S15).

#### 9.5 Ingredient Processing Score (IgFPro)

This section provides complementary information on the methods to create ingredient trees (Figure S16), and compares IgFPro with FPro (Figure S17).

### 10 Case Study on Rice Cakes

FPro can be leveraged to investigate the variations in the degree of processing within food subgroups and pinpoint different processing fingerprints. For example, the category of rice cakes comprises popular low-calorie carb snacks with a high glycemic index. In GroceryDB, there are 17 rice cakes displaying a wide range of variability in FPro (median *FPro* = 0.7964, first quartile Q1=0.6939, third quartile Q3=0.9302, see Figure S18). The top 3 processed rice cakes (“Great Value Birthday Cake Crispy Rice Treats, 0.78 oz, 8 Count”, “Quaker Gluten-Free Rice Cakes, Caramel, 6.5 Oz”, and “Quaker Chocolate Crunch Large Rice - Cakes 7.23oz”) have FPro values 0.9702, 0.9538, and 0.9372 respectively, reflecting the need to apply food processing to remove gluten (e.g., removing germ from corn as indicated on the product’s ingredient list) and the use of sorghum and corn syrups, colors, chocolate chips, resulting in higher sugar content. Conversely, among the least processed rice cakes we find “Quaker Lightly Salted Gluten Free Rice Cakes - 4.47oz” (*FPro* = 0.6802) and “Organic Cinnamon Toast Rice Cakes, 9.5 oz” (*FPro* = 0.6473) which exhibit simpler ingredients as well as lower sugar and the use of organic ingredients. Yet, from a broader perspective, none of these products can be considered “minimally-processed”, but rather “processed” or “ultra-processed”.

**Figure S13:**
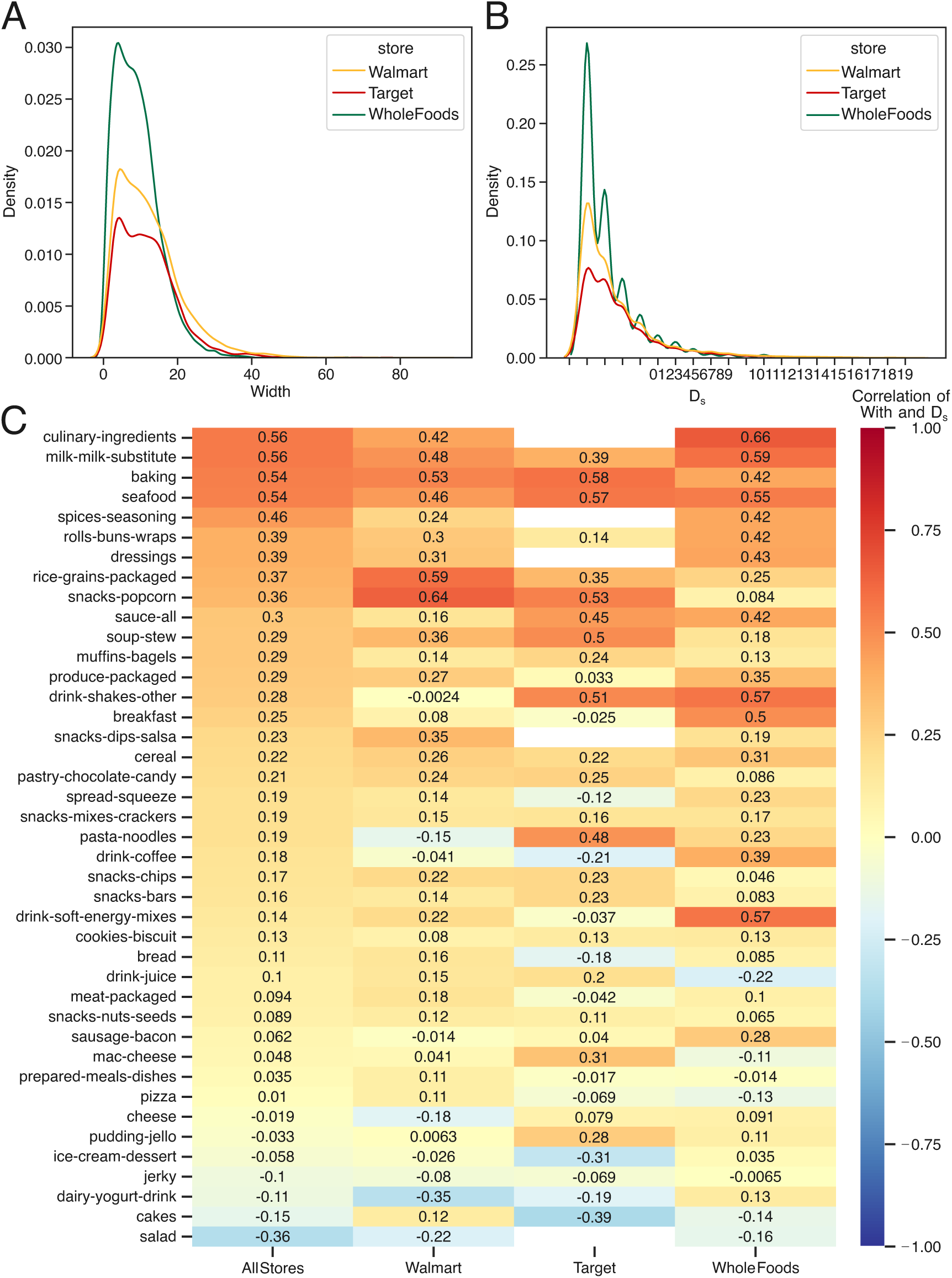
Characteristics of Ingredient Trees. **(a)** The distribution of the width of ingredient trees for each store, indicating that products in Whole Foods tends to have fewer ingredients compared to other stores. **(b)** The distribution of *D_s_* for all ingredient trees, reflecting the extent to which products rely on sub-ingredients. Whole Foods tends to rely less on mixing sub-ingredients compared to the other stores. **(c)** The Spearman’s correlation coefficient between the width and *D_s_* of ingredient trees for each harmonized category and store. Only products with an FPro scores were used in this analysis of tree depth and tree width.

**Figure S14:**
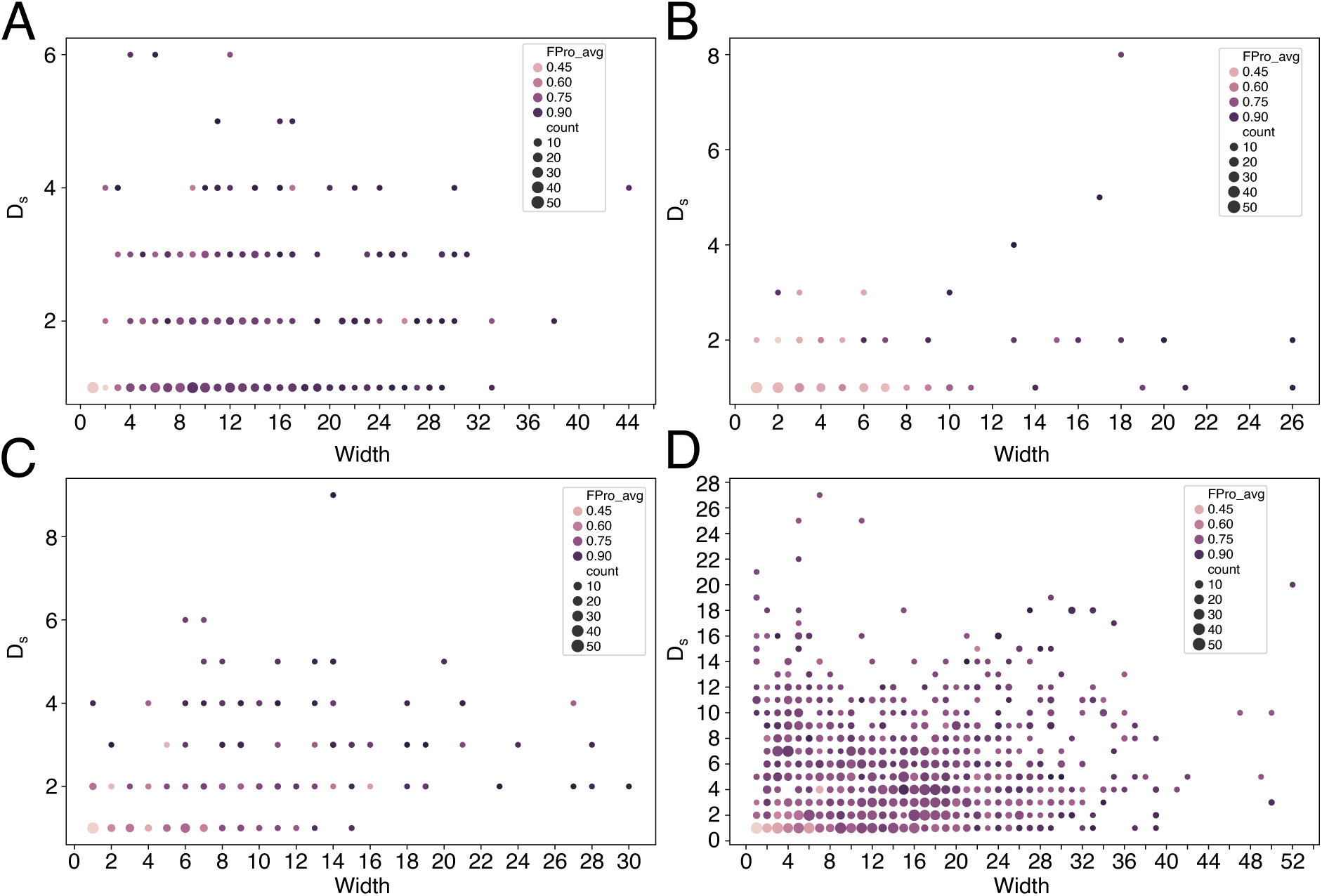
The Relationship between FPro and Characteristics of Ingredient Tree. Complex ingredient trees tend to have a higher FPro. **(A-C)** In cereals, pasta-noodles, and baking categories, there are FPro increases as tree width *W* and *D_s_* increases. **(D)** This effect is weaker in prepared meals & dishes.

**Figure S15:**
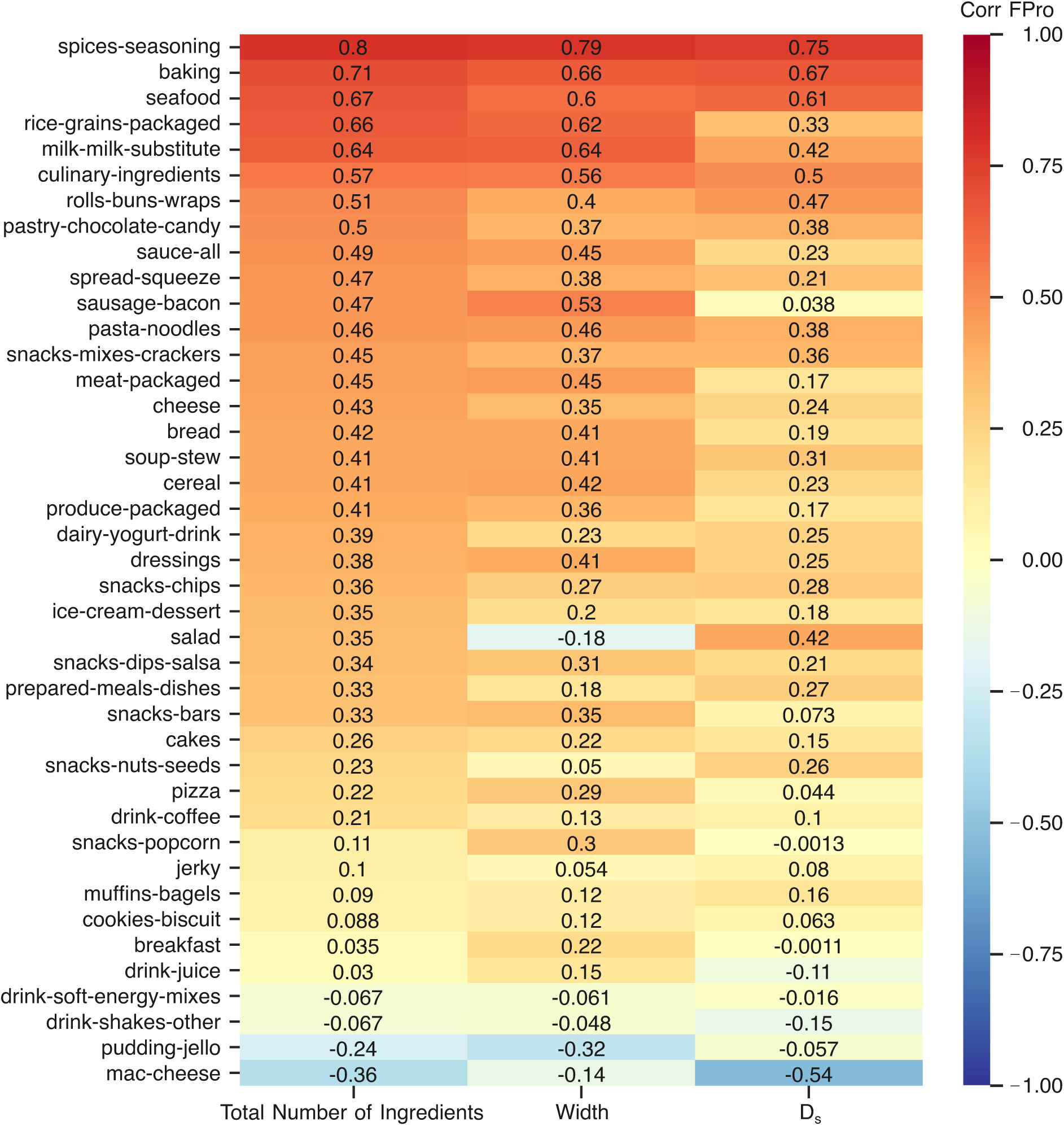
Correlation between FPro and the Characteristics of Ingredient Trees. The Spearman’s correlation between FPro and the ingredient trees width, *D_s_*, and total number of ingredients. The features of ingredient trees are not always positively correlated with FPro, depending on the food category. In some categories like seafood and milk-milk-substitute, there is a strong positive correlation between FPro and the characteristics of ingredient trees. However, negative correlations are observed in mac-cheese and pudding-jello.

**Figure S16:**
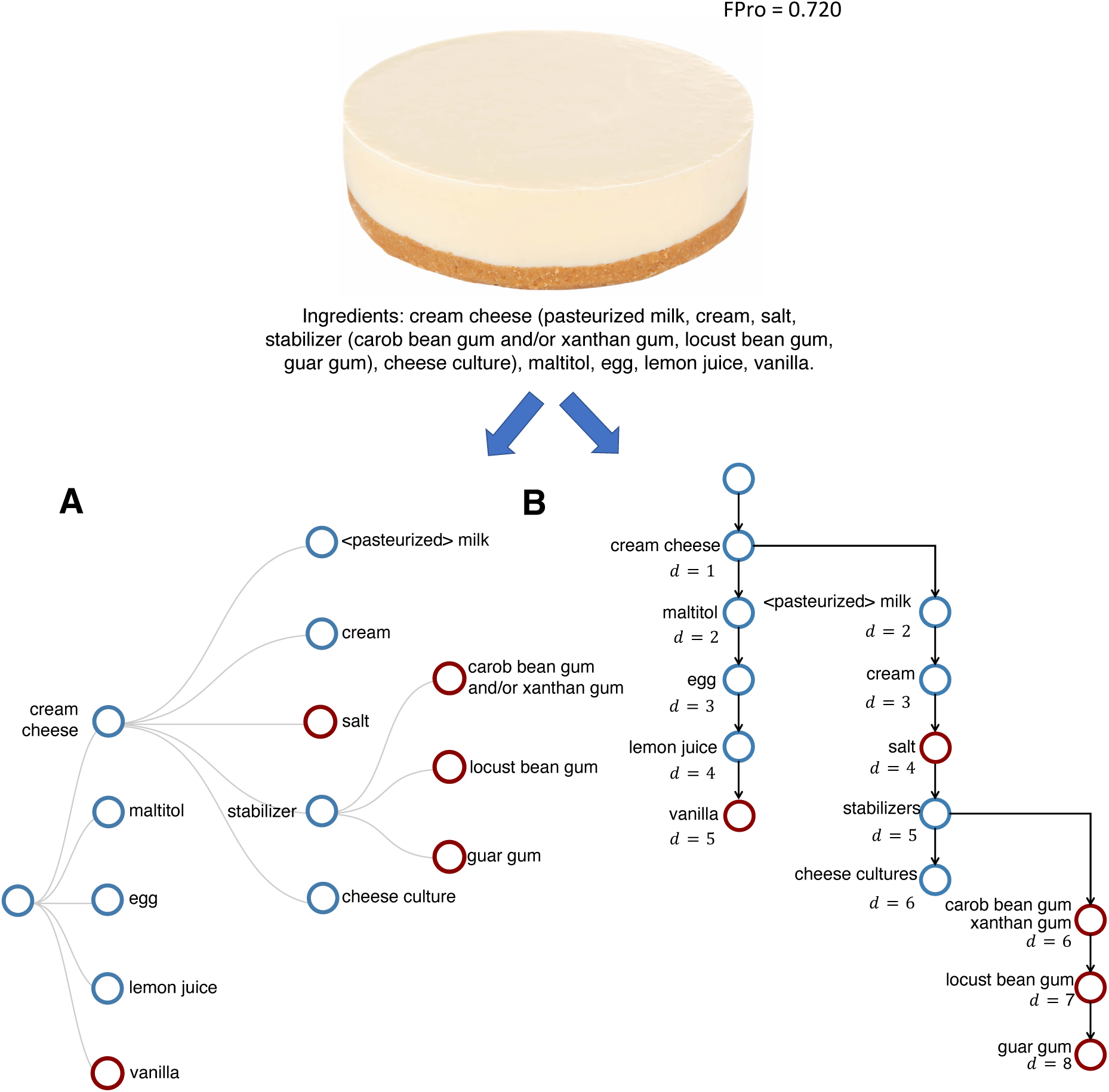
Two Types of Ingredient Trees. An ingredient list can be represented by two types of ingredient trees. **(a)** A recipe-like structure to better demonstrate the main and sub-ingredients. **(b)** A sequential approach to capture the order of ingredients in the tree structure. In this approach, the distance *d* from the root reflects a ranking for the amount of ingredients used in the preparation of an item. Image credits: “Delicious cheesecake on white background” (by Africa Studio for Adobe Stock).

List of all rice cakes in GroceryDB sorted by FPro (decreasing):

1. Great Value Birthday Cake Crispy Rice Treats, 0.78 oz, 8 Count (FPro: 0.9702) URL: [Product Link @ TrueFood.Tech]
2. Himalayan Sea Salt Sprouted Rice And Cauliflower Rice Cake, 5 oz (FPro: 0.9343) URL: [Product Link @ TrueFood.Tech]
3. Quaker Garden Tomato & Basil Rice Cakes - 6.1oz (FPro: 0.9261) URL: [Product Link @ TrueFood.Tech]
4. Quaker Gluten-Free Apple Cinnamon Rice Cakes, 7.04 Oz. (FPro: 0.9197) URL: [Product Link @ TrueFood.Tech]
5. Quaker Caramel Corn Gluten Free Rice Cakes - 6.56oz (FPro: 0.8846) URL: [Product Link @ TrueFood.Tech]
6. Quaker Rice Cakes, Apple Cinnamon, 6.53 Oz (FPro: 0.8256) URL: [Product Link @ TrueFood.Tech]
7. Quaker Rice Cakes, Lightly Salted, 4.47 Oz (FPro: 0.7964) URL: [Product Link @ TrueFood.Tech]
8. Organic Tamari With Seaweed Rice Cakes, 8.5 oz (FPro: 0.7858) URL: [Product Link @ TrueFood.Tech]
9. Organic Lightly Salted Brown Rice Cakes, 8.5 oz (FPro: 0.7562) URL: [Product Link @ TrueFood.Tech]
10. Organic Lightly Salted Wild Rice Cakes, 8.5 oz (FPro: 0.7562) URL: [Product Link @ TrueFood.Tech]
11. Rice Cakes Blueberry & Beet, 1.4 oz (FPro: 0.7076) URL: [Product Link @ True-Food.Tech]
12. Quaker Lightly Salted Gluten Free Rice Cakes - 4.47oz (FPro: 0.6802) URL: [Product Link @ TrueFood.Tech]
13. Organic Cinnamon Toast Rice Cakes, 9.5 oz (FPro: 0.6473) URL: [Product Link @ TrueFood.Tech]
14. Quaker Large Rice Cake Apple Cinn - 6.53oz (FPro: 0.6365) URL: [Product Link @ TrueFood.Tech]
15. Organic Salt Free Brown Rice Cakes, 8.5 oz (FPro: 0.5182) URL: [Product Link @ TrueFood.Tech]

**Figure S17:**
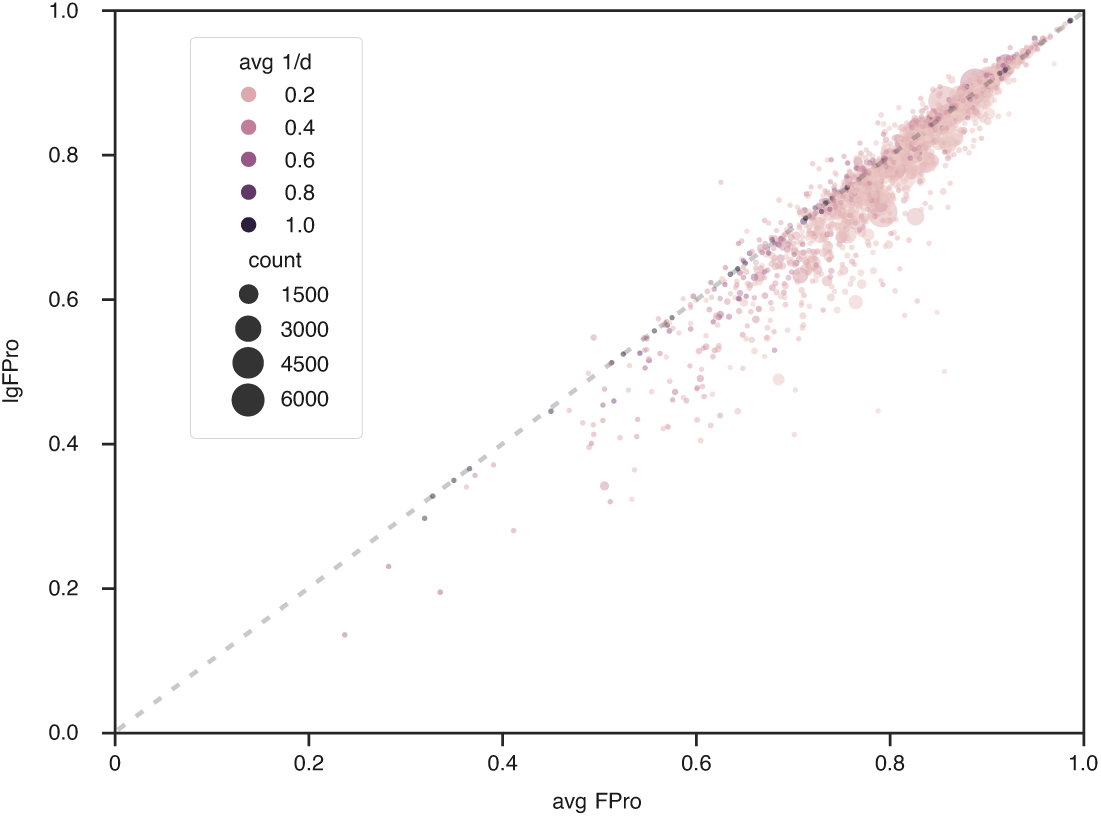
IgFPro vs ⟨FPro⟩. IgFPro shows remarkable variability when compared to the average FPro of products containing the selected ingredient, suggesting distinctive patterns of correlation between the products’ FPro and the ranking of ingredients in their ingredient lists. In this plot, the general form of ingredients is used without descriptors. For example, the general name of ‘milk *<*pasteurized*>*’ without descriptors is ‘milk.’ Also, only the general ingredients that are present in at least 10 products are considered, resulting in IgFPro measures for over 1,200 ingredients, as ranked in Figure 6.

**Figure S18:**
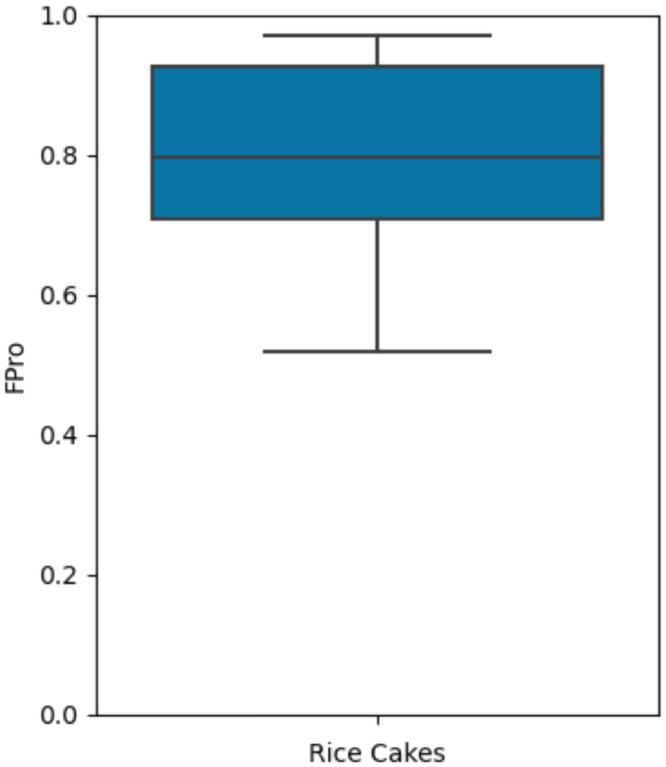
Distribution of FPro for all rice cakes in GroceryDB (total 17). For the box plots, the minimum is the lower quartile, the central line is the median, and the maximum is the upper quartile. The whiskers show data outside of the inter-quartile range. Diamonds represent outliers.

## Notes

### Summary of Updates

We updated the parsing and the disambiguation of the ingredient lists

